## Supplementary Materials for "The shape of cancer relapse: Topological data analysis predicts recurrence in paediatric acute lymphoblastic leukaemia"

**This PDF file includes:**

Supplementary Materials Section 1: Topological data analysis and persistent homology

Supplementary Materials Section 2: Methodology structure

**Other Supplementary Materials for this manuscript include the following:**

Figs. S1 to S23

Tables S1 to S8

Data S1 to S3

### Supplementary Materials Contents

**1 Topological data analysis and persistent homology** **3**

### 1 Topological data analysis and persistent homology

Flow cytometry data, as many other medical datasets, is high-dimensional, i.e. the number of immunophenotypical markers and the number of cells analysed is very high (on the order of millions). Currently, state-of the art analyses and diagnostic decisions are based on estimating the shape of projections of the data to two or three dimensions [12]. Not only does this result in a loss of information in the data, but it also leads to subjective and unreproducible decisions made by individual doctors. Novel methods are needed to obtain objective summary features from these high-dimensional data, ideally in a fully automated, quick, and intuitive manner.

An emerging mathematical field that uses topological and geometric approaches to quantify the "shape" of data in a completely objective and reproducible manner is *topological data analysis (TDA)* [22,23] (for excellent informal introductions see also [75-77] ). TDA is not limited to projections, but can take into account the full high-dimensional structure of the data. The input to algorithms from TDA is data, for example in the form of a point cloud, i.e. coordinates in space. This point cloud is then equipped with a mathematical structure, a so-called simplicial complex, which allows its topological analysis. The shape of the data is then characterised via topological invariants, for example connected components and loops formed by data points surrounding an empty hole. A central method in TDA is *persistent homology* (PH) [22-26] . PH considers topological invariants at different spacial scales. In general, invariants that persist over many such scales are considered to be more representative of the data than invariants that only appear over a small range of scales.

Improved computational feasibility of PH [26]has increased its applications to (high-dimensional) biomedical data. For example, such applications include studies of brain arteries [30], neurons [31], airways [33], stenosis [34], zebrafish patterns [35], contagion dynamics [36], blood vessel networks of tumours [40,78], and spatial networks [39,41,42]. PH has also been successfully applied to classify synthetic data from mathematical models of angiogenesis, the process in which tumour blood vessels form from existing ones [44].

#### 1.1 Simplices and simplicial complexes

A topological space can be approximated by structures called simplicial complexes. These can be thought of as generalised graphs which are composed of simplices in various dimensions: points (0-simplices), edges (1-simplices), triangles (2-simplices), tetrahedra (3-simplices) (see Figure S1(A)). The boundary of higher-dimensional simplices are lower-dimensional simplices. For example, a triangle has three edges forming its boundary, an edge has two points as its boundary. The boundary map is a linear map which sends a simplex to the simplices on its boundary (see Figure S1(B)). A simplicial complex is constructed by glueing together simplices from various dimensions along a lower-dimensional simplex.

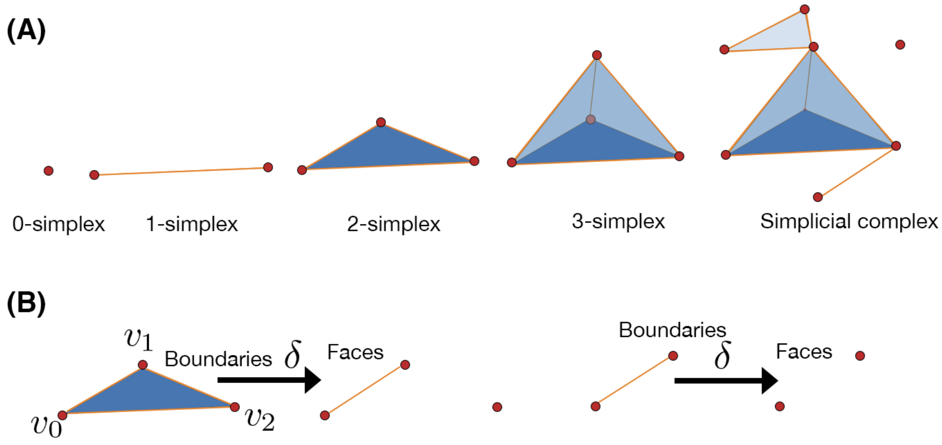

Figure S1: **Basics of simplicial complexes.(A)** Topological spaces can be simplified by simplices (points, edges, triangles, tetrahedra etc.), which can be glued together in simplicial complexes. **(B)** By taking the boundary map of a simplex, one can mao it to the simplices on its boundary.

#### 1.2 Homology

Homology is a way of counting connected components or loops in simplicial complexes. Intuitively, homology in a set dimension is a lense through which one can view the simplicial complex to visualise the loops in this dimension. For example, in dimension 1, homology highlights sequences of pairwise connected points, i.e. edges, forming a loop, but omitting any edges that are boundaries of higher dimensional simplices, e.g. edges that are boundaries of a triangle.

#### 1.3 Persistent homology and barcodes

In order to apply the theory of homology to point cloud data, one first needs to connect the data points to form a simplicial complex. One possibility is the Vietoris-Rips complex [71]: for a given radius $r$, we draw a ball centred at each data point. If two such balls intersect, we connect the data points with an edge, i.e. a 1-simplex. If three data points are connected pairwise to form a triangle, we consider this to be a 2-simplex. We continue analogously for higher dimensions (see S2(A)). An appropriate choice of radius is, however, not clear a priori. For example, if the radius is too small, no points are connected; if the radius is too big, all points are connected by edges and no holes are visible (see Figure S2(B)). Persistent homology therefore considers a whole sequence of increasing radii, which give rise to a sequence of embedded simplicial complexes, a so-called filtration. Note that it is possible to create filtrations from data in many different ways and the choice depends on the problem at hand [77]. In our case, we use the Vietoris-Rips filtration.

We can track topological features such as connected components and loops along a filtration. Intuitively, when increasing the radius $r$ of balls centred at data points, topological features appear at a "time" $r_{\text{birth}}$ and disappear at a time $r_{\text{death}}$. This leads to a definition of "lifetime" $\rho_r = r_{\text{death}} - r_{\text{birth}}$, or persistence. Persistence can be visualised by barcodes, where persistence

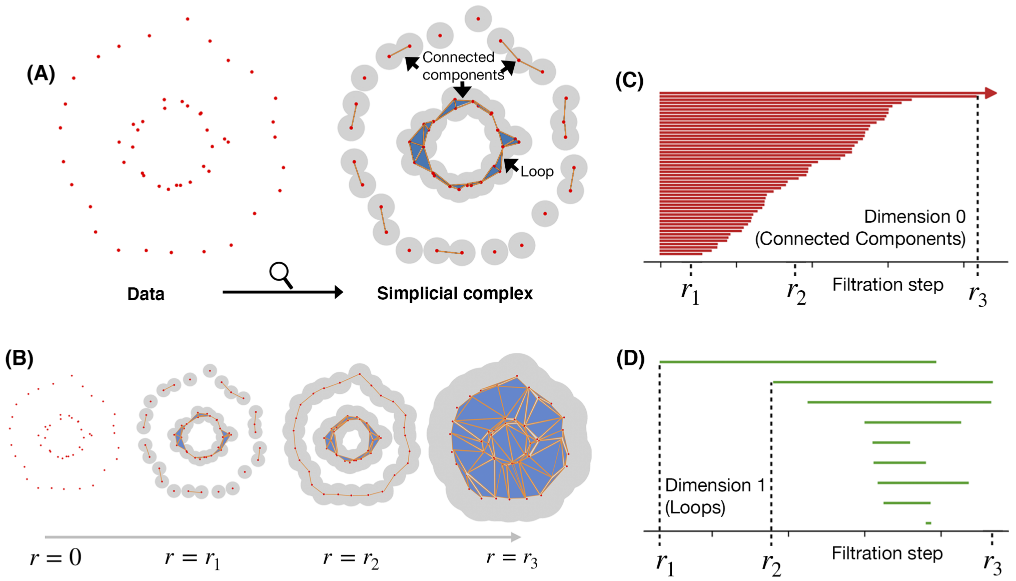

Figure S2: **Basics of persistent homology. (A)** By defining a filtration of simplicial complexes on point cloud data we can study how loops and connected components are formed and change over different spatial scales. **(B)** When increasing the radius $r$ of balls centred at data points, topological features such as connected components and loops arise: At $r = r_1$, a loop appears, followed by a second one at $r = r_2$. For $r = r_3$, all points are connected in the same connected component. **(C)** Persistence barcode for dimension 0 of data in (A). Each bar represents a topological feature and its lifetime, i.e. persistence, in the filtration. The top bar is the only one that persists longer than $r_3$ and indeed persists until the end of the filtration. This is also referred to as infinite persistence. **(D)** Persistence barcode for dimension 1 of data in (A). Each bar represents a loop. The longest persisting loops are "born" at $r = r_1$ and $r = r_2$.

intervals $[r_{\text{birth}}, r_{\text{death}})$ are represented, see Figure S2(C) and (D) for dimension 0 and dimension 1, respectively.

#### 1.4 Persistence diagrams and persistence images

An alternative representation of the output of PH is a persistence diagram. Similar to a barcode, it consists of a collection of radius pairs that correspond to the birth and death radii of topological features in the filtration, but in this case they are represented as coordinates $(r_{\text{birth}}, r_{\text{death}})$ rather than intervals (see Figure S3(A)). In persistence diagrams, points close to the diagonal represent short-lived features which are commonly interpreted as noise.

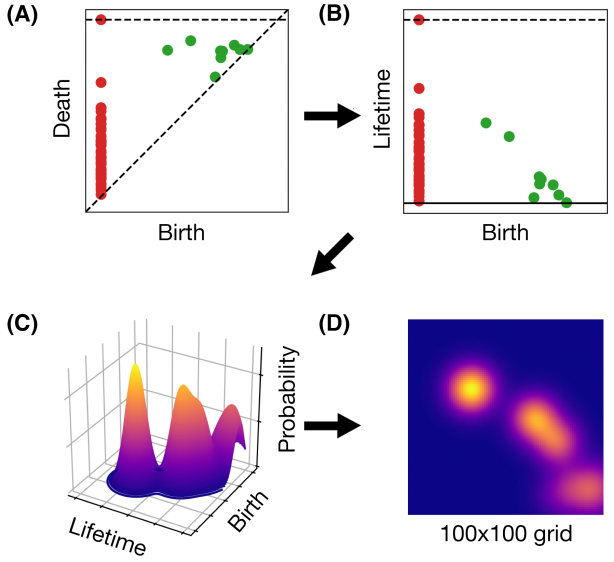

Figure S3: **Pipeline for Persistence Images**. **(A)** Persistence diagrams are constructed using the same information as in persistence barcodes, e.g. those in Figure S2(C) and (D), but interpreting the birth and death radius pairs as points in the plane rather than intervals. Points close to the diagonal represent topological features with short persistence while the horizontal line represents the end of the filtration and thus points on this line persist infinitely. We compute PH for dimension 0 (red points), and 1 (green points) and summarise the PH output in the same persistence diagram. **(B)** To obtain a persistence image, the first step is to consider the persistence diagram in birth-lifetime coordinates $(r_{\text{birth}}, r_{\text{death}} - r_{\text{birth}})$. **(C)** We generate 2D Gaussian distributions centered around each point, in this case shown for dimension 1 of the persistence diagram (B), and obtain a surface. **(D)** We then sum the volume within the surface (C) over a grid with a set pixel resolution discretising the birth-lifetime axes, here 100 x 100, and obtain a persistence image.

In our study, we worked with persistence images, generated from persistence barcodes or diagrams. These images can be viewed as real-valued matrices which, like barcodes, are stable under small perturbations of the input [54]and can be used as input into a variety of machine-learning and classification algorithms. We constructed persistence images as described in [54]using 2D Gaussian distributions with variances 0.01 and 0.05 and a grid size of 50$\times$50 and 100$\times$100 pixels. We show the pipeline as well as a typical persistence image in Figure S3). Persistence images have been successfully applied in a biological context in [44,80].

### 2 Methodology structure

In this work, we analysed flow cytometry data from relapsing (R) and non-relapsing (NR) patients with persistent homology. We now summarise the main conclusions and results obtained in our work:

#### 2.1 Data processing

We gathered both clinical and bone marrow data from patients diagnosed with acute lymphoblastic leukaemia. The clinical information can be found in Table S1. The bone marrow, quantitative data from the patients were obtained via flow cytometry. This technique is able to identify marker expression at a single-cell level.

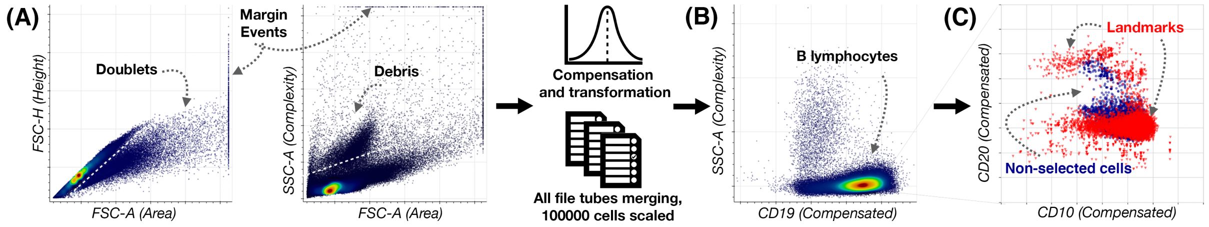

Figure S4: **Pipeline used to preprocess flow cytometry data. (A)** Manual gating was used to select B lymphocytes. First, doublets (i.e., two or more cells considered as single events) were removed from the panel FSC-A and FSC-H, below the white dashed line. Secondly, debris was removed from the panel FSC-A and SSC-A, omitting highly complex cells over the white dashed line. In both panels, events at the margins (located at the extreme of the axes) are removed. Next, single files were compensated and transformed, and finally merged into one file per patient. $10^5$ cells were taken randomly and scaled to avoid outliers. **(B)** We obtained the B lymphocytes by selecting $CD19^+$ cells. **(C)** The max-min algorithm was applied to obtain landmarks from the B lymphocyte point cloud. Using $10^4$ selected landmarks gave us a plausible representation of the whole B lymphocyte dataset.

##### 2.1.a Preprocessing, biomedical steps

Firstly, all patients samples were inspected manually by standard methods (see Figure S4). The data from the bone marrow was transformed into files with tables, where each cell has a certain level of intensity depending on the markers included in each sample. All samples files from each patient were compensated and transformed, and then merged into a single file. Therefore, we obtained a single file per patient with the following 16 common parameters: FSC-A, SSC-A, CD10, CD13, CD19, CD20, CD22, CD3, CD33, CD34, CD38, CD45, CD58, CD66c, IGM, cyCD3, cyMPO, cyTDT.

##### 2.1.b Division in discovery and validation set

Patients were analysed considering their relapse status (80 R, 16 NR). We divided our patients dataset into a discovery (Dataset 1 and 2) and a validation group (Dataset 3).

| | Dataset 1 (HVR) (N=30) | Dataset 2 (HNJ) (N=20) | Dataset 3* (HVA) (N=46) | Total (N=96) |
|---|---|---|---|---|
| Sex - no. (%) | | | | |
| Male | 19 (63) | 10 (50) | 23 (50) | 52 (54) |
| Female | 11 (37) | 10 (50) | 23 (50) | 44 (46) |
| Age at diagnosis - yr | | | | |
| Median | 2.5 | 4 | 4.5 | 4 |
| Range | 0 - 12 | 0-16 | 0 - 13 | 0-16 |
| Long term status -no. (%) | | | | |
| Relapse | 8 (27) | 5 (25) | 3 (7) | 16 (17) |
| No relapse | 22 (73) | 15 (75) | 43 (93) | 80 (83) |
| Immunophenotype - no. (%) | | | | |
| Common | 19 (64) | 10 (50) | 35 (76) | 64 (67) |
| Pre-B | 9 (30) | 2 (10) | 8 (17) | 19 (20) |
| Pro-B | 1 (3) | 1 (5) | 3 (7) | 5 (5) |
| Mixed | 1 (3) | 7 (35) | 0 (0) | 8 (8) |
| BM blasts at diagnosis - % | | | | |
| Median | 81.95 | 87.5 | 79.4 | 81.6 |
| Range | 25.0-96.3 | 62.0-99.0 | 25.6 - 99.0 | 25.0-99.0 |
| Risk at diagnosis - no. (%) | | | | |
| High | 0 (0) | 0 (0) | 6 (13) | 6 (6) |
| Intermediate | 12 (40) | 15 (75) | 18 (39) | 45 (47) |
| Low | 17 (57) | 2 (10) | 22 (48) | 41 (42) |
| Not available | 1 (3) | 3 (15) | 0 (0) | 4 (5) |
| Karyotype - no. (%) | | | | |
| Hyperdiploid (>50) | 10 (33) | 5 (25) | 2 (4) | 17 (18) |
| Normal (40-50) | 10 (33) | 15 (75) | 9 (20) | 34 (35) |
| Hypodiploid (<40) | 0 (0) | 0 (0) | 0 (0) | 0 (0) |
| No metaphases | 10 (33) | 0 (0) | 35 (76) | 45 (47) |
| Chromosomic alterations - no. (%) | | | | |
| ETV6/RUNX1 t(12;21) | 4 (13) | 2 (10) | 10 (22) | 16 (17) |
| TCF3/PBX1 t(1;19) | 0 (0) | 1 (5) | 0 (0) | 1 (1) |
| MLL rearrangement | 3 (10) | 0 (0) | 2 (4) | 13 (5) |
| BCR/ABL1 t(9;22) | 0 (0) | 0 (0) | 0 (0) | 0 (0) |

* In HVA, one patient from HVA lacked data on MLL rearrangement and 2 patients from BCR/ABL1 t(9;22).

Table S1: **Patients clinical features**. Datasets 1 and 2, from Hospital Niño Jesús (HNJ) and Hospital de Vírgen del Rocío (HVR) correspond to the validation group. Dataset 3, from Hospital Virgen de la Arrixaca (HVA) corresponds to the test group. Most features measure the number of patients (N) and its percentage (in brackets). For the age at diagnosis as well as the percentage of bone marrow (BM) blasts at diagnosis, median and range parameters are included.

##### 2.1.c Previous clinical analysis

We checked for differences between the risk assessment based on clinical decision (low, intermediate and high) and the R/NR status of each patient (see Data S4 for details). This summary can be found in Table S2.

| **Long term status** | **Risk at diagnosis** | Percentage (Count) |
|---|---|---|
| No relapse | High | 5.2% (4) |
| | Intermediate | 47.4% (37) |
| | Low | 47.4% (37) |
| Relapse | High | 14.3% (2) |
| | Intermediate | 57.1% (8) |
| | Low | 28.6% (4) |

Table S2: Frequencies and count of patients with data on risk stratification (92 out of 96) at the moment of diagnosis in terms of their long term status.

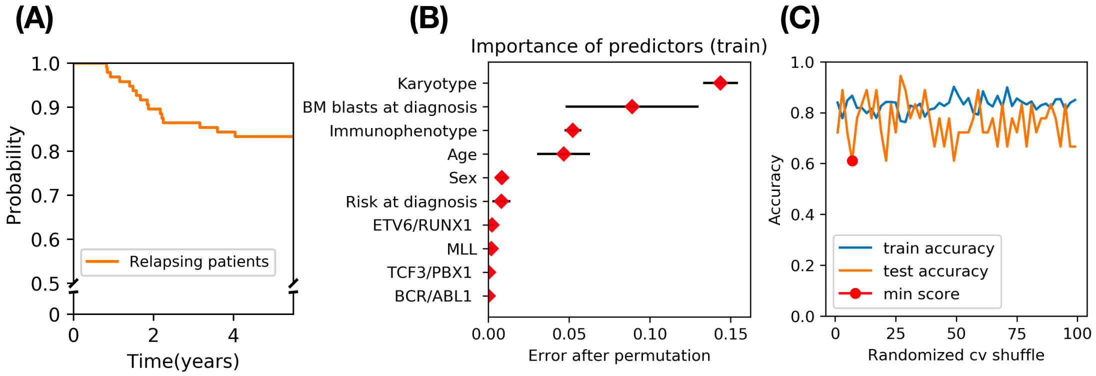

Figure S5: **Description and results on clinical variable analysis.** **(A)** Kaplan Meier curve on patients' relapse times. **(B)** Classification in Random forest showed that most important features were karyotype, number of bone marrow (BM) blasts at diagnosis, immunophenotype and age. **(C)** Accuracy results on classification based on the previous variables for training and test sets depending on each training set fromo the shuffle obtained in cross-validation (cv).

As seen in Table S2, 4 R patients were assigned a low risk at diagnosis, and 4 NR patients were assigned a high risk at diagnosis. In these terms, only 8 patients out of the 92 with clinical information would be misclassified (accuracy of 91.3%). Patients experience relapse up to 4 years from the moment of diagnosis, as shown in Figure S5. We classified the patients in terms of the R/NR status considering the clinical variables from Table S1. We performed several Random Forest analyses on the discovery and validation set without empty values on the clinical data (8 R, 38 NR patients). To do so, we randomly shuffled training and test sets (cross-validation). After permutating the predictors used, the variables karyotype, number of bone marrow (BM) blasts at diagnosis, immunophenotype and age were the most important in train sets. The results showed a mean accuracy of 86% in the classification of training sets and a 75% for test sets, with a minimal classification accuracy of 52.3%.

#### 2.2 Persistent homology analysis: obtention of biomarkers

##### 2.2.a Persistence barcodes of all pairwise combinations of potential biomarkers

To obtain a set of candidate biomarkers, we considered pairwise combinations of all 16 immunophenotypic (IPT) markers and computed their persistence barcodes. Then, we computed simple barcode summaries: the maximum, minimum, standard deviation, and mean persistence in dimensions 0 and 1. We classified our patients considering their R or NR status using these simple barcode summaries as input feature vectors into a Random Forest. The classification results for the whole dataset performing oversampling can be found in Data S1. The results using stratified k-fold for the validation and test group can be found in Data S1 and Data S2. To check for possible correlation between the markers, an analysis in the R and NR sets are shown respectively, in Figures S6 and S7.

##### 2.2.b Selection of biomarkers with best area under the curve

From the classification results, we concluded that both in the validation and test set, only combinations including CD10, CD20, CD38 and CD45 had an area under the curve (AUC) greater than 50%.

We performed a statistical analysis based on maximal, minimal, mean, and standard deviation of the persistence features for the CD10-CD20-CD38-CD45 biomarkers in four dimensions for relapsed and non-relapsed patients. We include results of these analyses in Figure S8 (A). However, there were no significant features when comparing R and NR patients, as the p-values were larger than 0.05 in the discovery set.

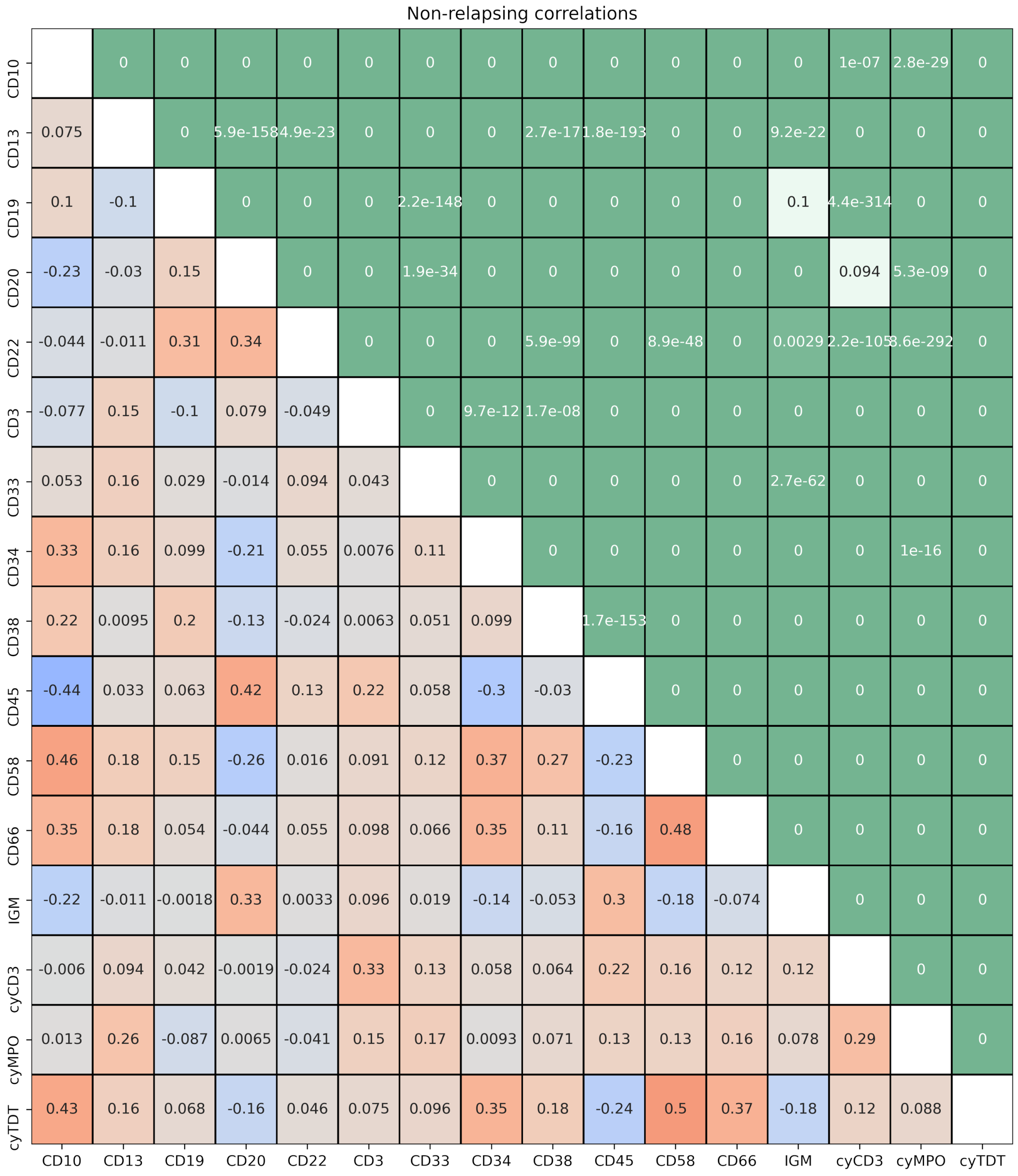

Figure S6: **Correlation of markers in NR patients.** The p-value is shown in the top triangle, while in the bottom triangle the correlation rate is shown in a scale from direct (orange) to inverse correlation (blue). No strong correlations are present between the variables.

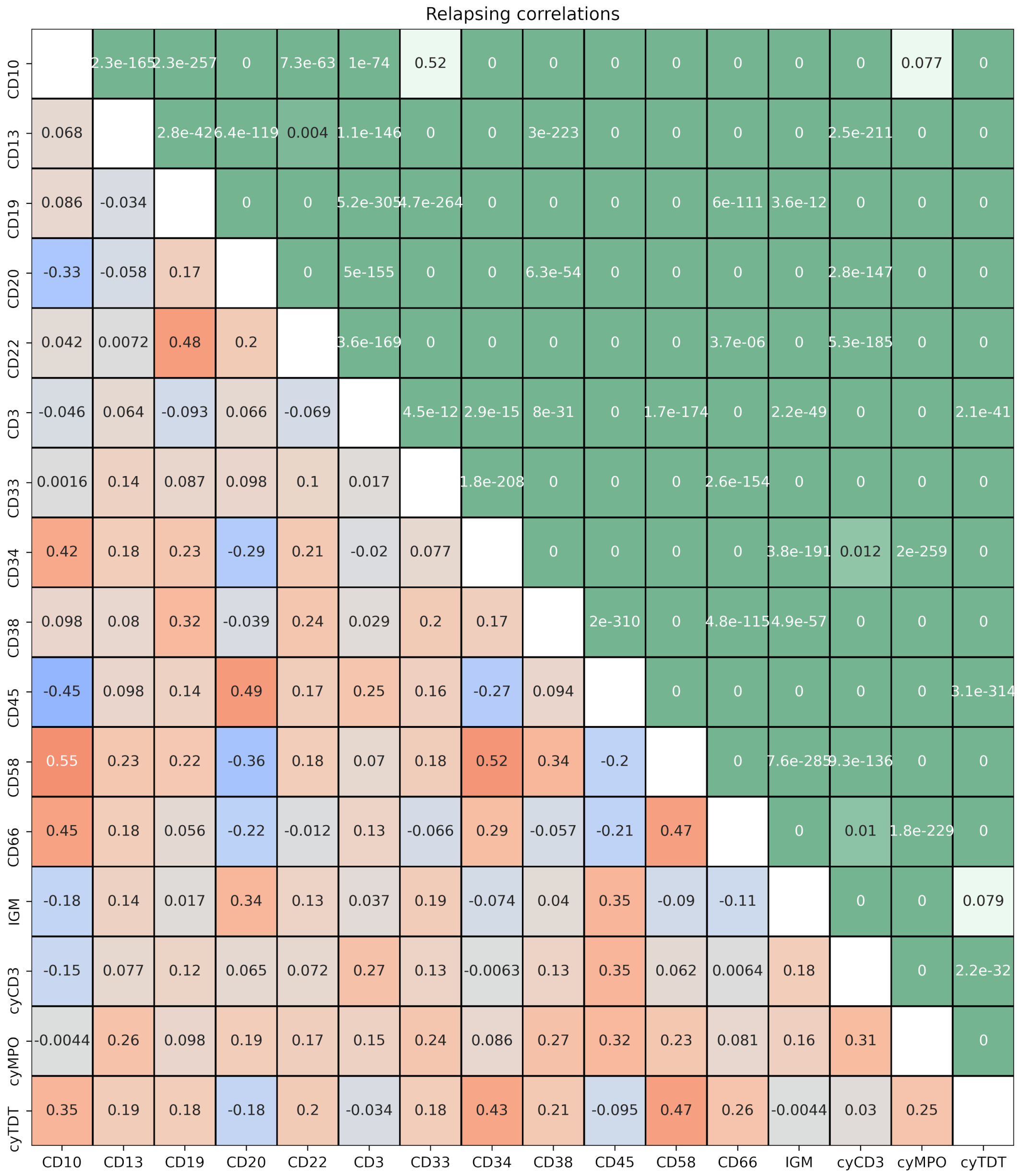

Figure S7: **Correlation of markers in R patients.** The p-value is shown in the top triangle, while in the bottom triangle the correlation rate is shown in a scale from direct (orange) to inverse correlation (blue). No strong correlations are present between the variables.

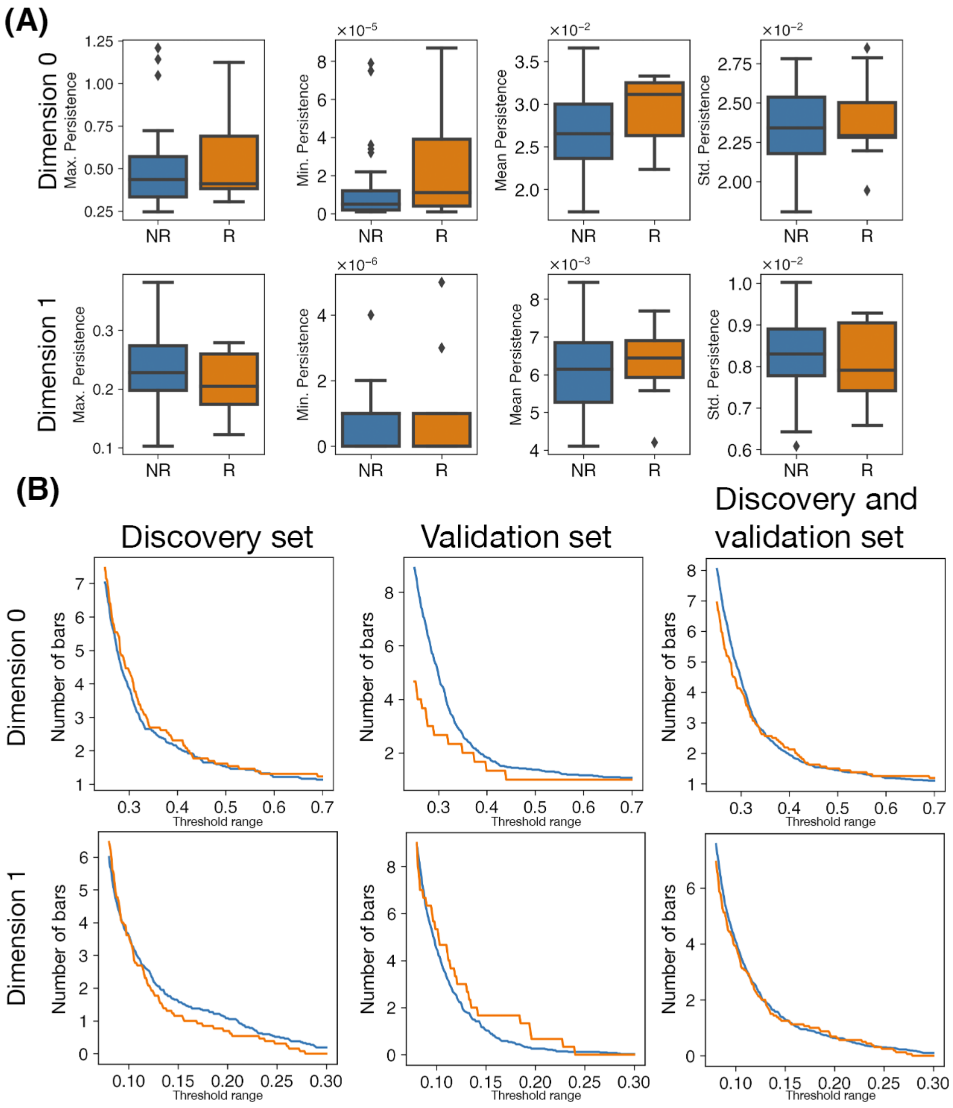

Figure S8: **Persistence analysis of four-dimensional space of CD10-CD20-CD38-CD45 showed topological differences between R (orange) and NR patients (blue)**. **(A)** Boxplots of maximal, minimal, mean, and standard deviation persistence of non-relapsed (blue) and relapsed patient data (orange). **(B)** Difference in PT curves in markers CD10-CD20-CD38-CD45 for R (orange lines) and NR patients (blue lines) for set ranges of filtration steps, for the discovery set and validation set. We found discrepancies between the discovery and validation dataset for dimension 0 (connected components), in the threshold range $\tau \in [0.2, 0.7]$. For dimension 1, we found statistical significance but with a higher number of loops in NR patients for the discovery set, and conversely in the validation set for $\tau \in [0.05, 0.30]$. Considering both datasets together, we found no difference as there was no statistical significance between both groups of patients.

#### 2.3 Interpretation and classification based on the biomarkers

##### 2.3.a 4-dimensional analysis of CD10-CD20-CD38-CD45

We further exploited more information from the PH barcodes and computed how the number of connected components (dimension 0 for connected components) and the number of loops (dimension 1 for loops) change along the different spatial scales of PH. To do so, we constructed the so-called Persistence Threshold (PT) curves, by counting the number of bars in each barcode whose corresponding topological features persist longer than a prescribed range of set thresholds. Their changes along the spatial range of these thresholds give rise to PT curves. Thus, we checked the difference between the persistence threshold curves between the R and NR groups in markers CD10-CD20-CD38-CD45. This was performed in dimension 0 and 1 and for certain threshold ranges, for the discovery set and validation set (see Figure S8 (B)). We found discrepancies between the discovery and validation dataset for dimension 0 (connected components), in the threshold range $\tau \in [0.2, 0.7]$. For dimension 1, we found statistical significance but with a higher number of loops in NR patients for the discovery set, and conversely in the validation set for $\tau \in [0.05, 0.30]$. Considering both datasets together, we found no difference as there was no statistical significance between both groups of patients. These differences showed discrepancies, for which we recurred to a 2-dimensional analysis of the pairwise combinations of the biomarkers.

##### 2.3.b 2-dimensional analysis of pairwise combinations of CD10-CD20-CD38-CD45

We performed PH of the pairwise combination of biomarkers CD10-CD20-CD38-CD45. We computed the number of bars longer than a set of thresholds in dimensions 0 and 1 for R and NR patients in Figures S10 and S12, and for the normalised number of bars (i.e. the percentage over the total amount of bars in each patient) in Figures S11 and S13. This analysis was performed in the discovery and validation sets. As observed, several combinations were in agreement between discovery and validation set, with p-values were below 0.05 as presented in Figures S14 and S15. For the combinations CD10-CD20, in dimension 0, R patients presented a higher number of connected components (see Figure S9) for all thresholds. For the combinations including CD10 or CD20, R patients also presented a higher number of loops, while for the combination CD38-CD45, NR patients presented a higher number of loops.

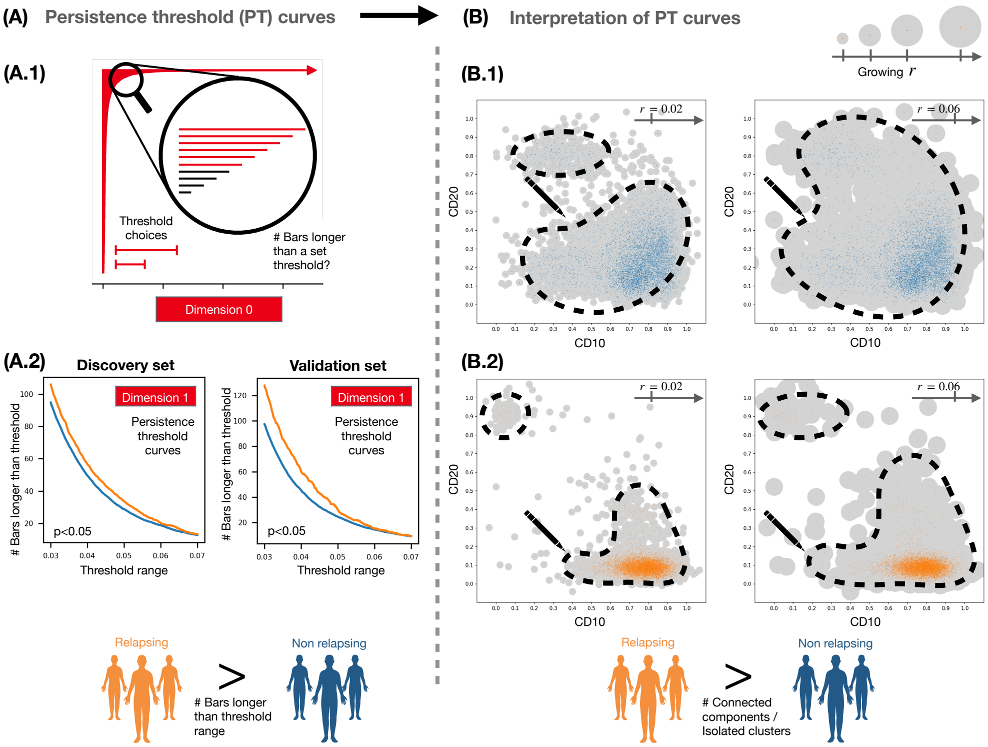

Figure S9: **Analysis of persistent homology output and interpretation of results.** **(A)** We computed the number of bars in dimension 0 persistence barcodes that are longer than a set of threshold choices. (A.1). This provides information on the persistence of connected components in the data. In both the discovery and validation sets, R and NR patients differed statistically significantly in the number of bars for all pairwise combinations of markers CD10, CD20, CD38 and CD45 except for the case of CD10-CD45. These 'persistence threshold curves' demonstrated a higher number of persistent connected components and loops for R patients in comparison to NR patients. **(B)** As a consequence of our quantitative approach, we suggest a qualitative approach on the 2D projection CD10-CD20: consider balls of growing radii centred at each data point; our results indicate that individual connected components and loops merge quickly with one another as the radius grows for NR patients (B.1), while the connected components and loops for R patients remain separate for a longer range of radii (B.2).

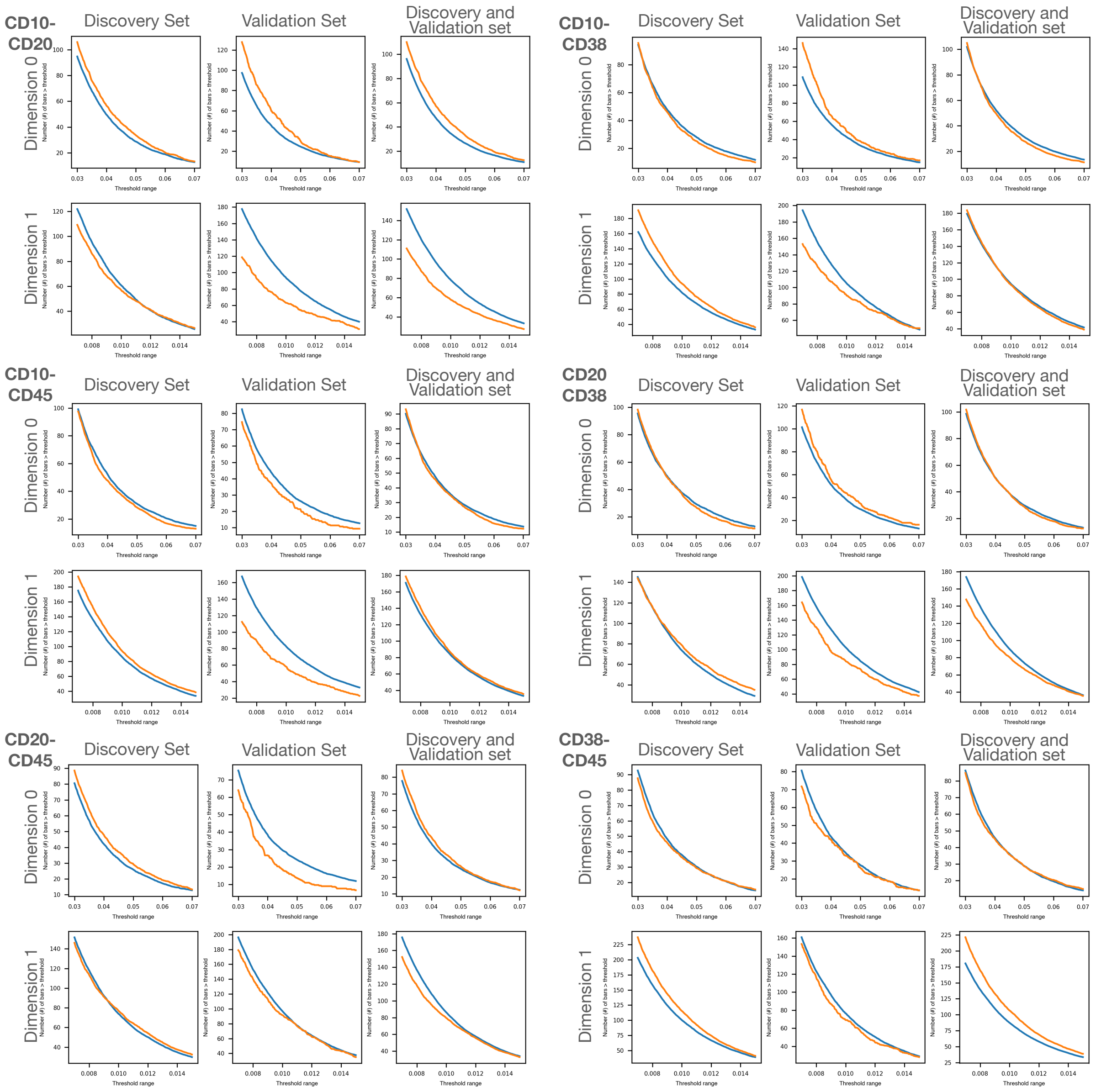

Figure S10: **Difference in the number (#) of bars with medium persistence in dimension 0 and 1 for the pairwise combination of markers CD10, CD20, CD38 and CD45.** R (orange lines) and NR patients (blue lines) PT curves are shown for set threshold ranges ($\tau \in [0.03, 0.07]$ for dimension 0 and $\tau \in [0.007, 0.015]$), for the discovery, validation, and both sets combines. P-values and value of the estimators after a t-test comparison are included in Figure S14.

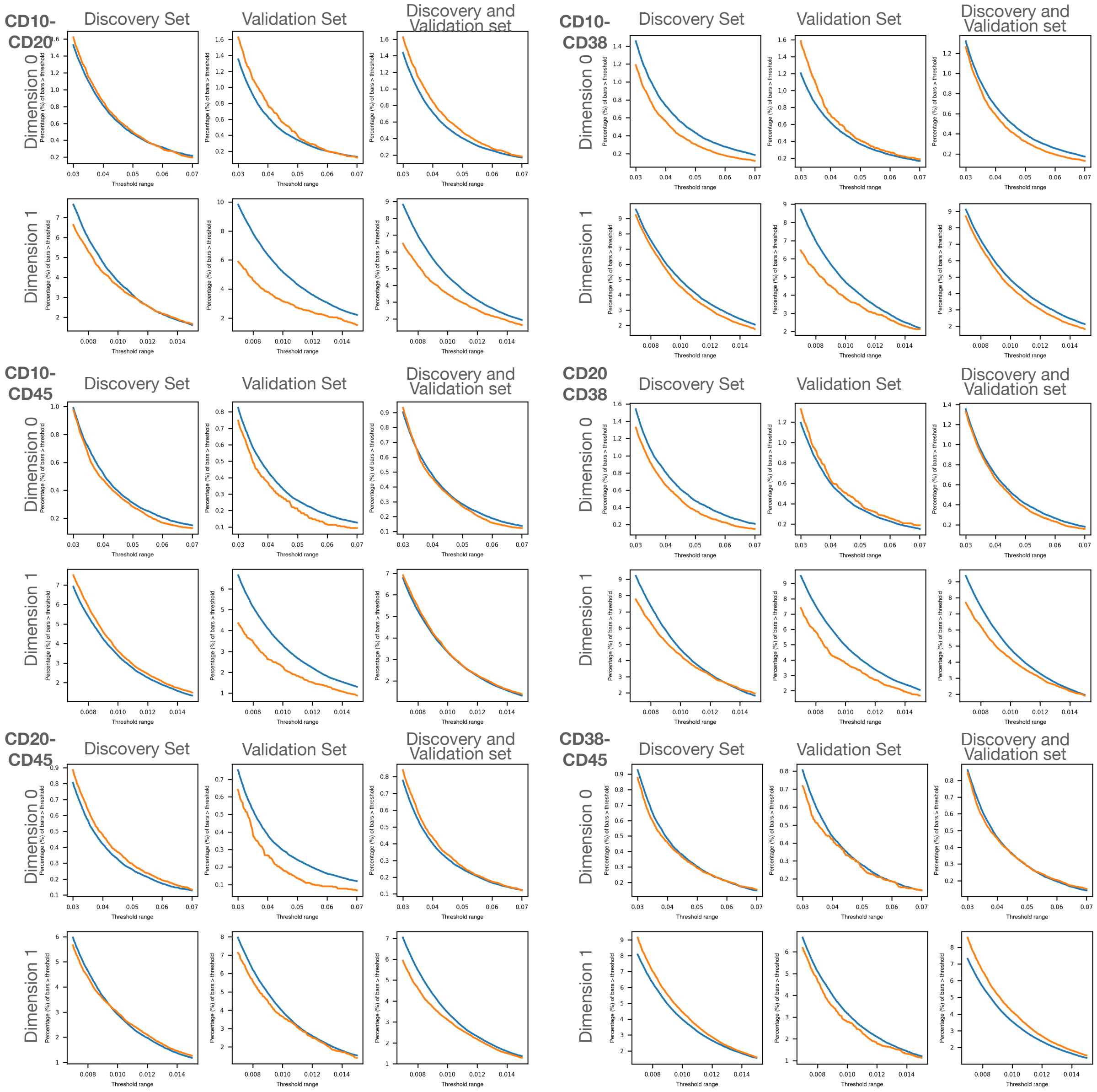

Figure S11: **Difference in the percentage (%) of bars with medium persistence in dimension 0 and 1 for the pairwise combination of markers CD10, CD20, CD38 and CD45.** R (orange lines) and NR patients (blue lines) PT curves are shown for set threshold ranges ($\tau \in [0.03, 0.07]$ for dimension 0 and $\tau \in [0.007, 0.015]$), for the discovery, validation, and both sets combines. P-values and value of the estimators after a t-test comparison are included in Figure S15.

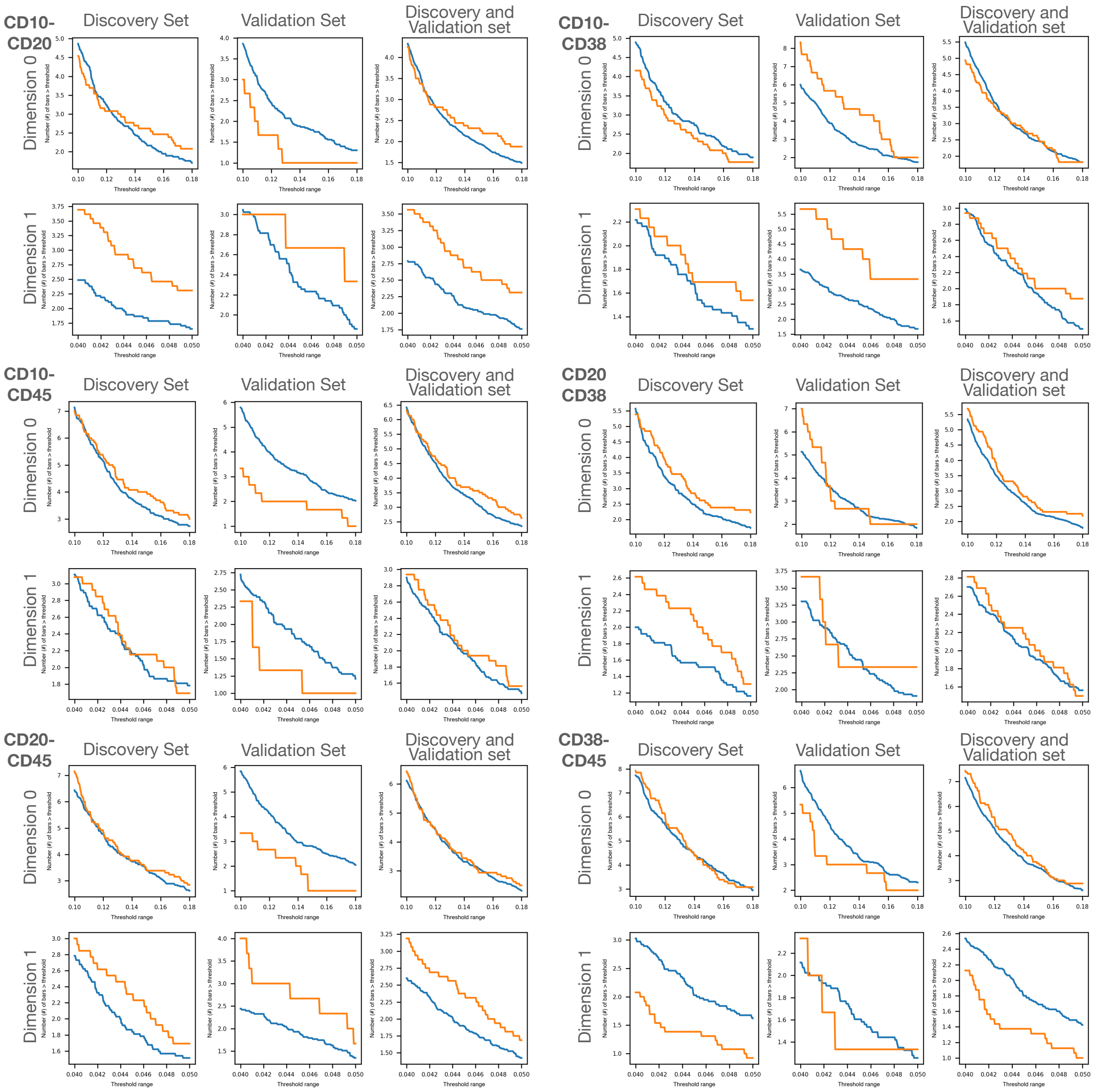

Figure S12: **Difference in the number (#) of bars with long persistence in dimension 0 and 1 for the pairwise combination of markers CD10, CD20, CD38 and CD45.** R (orange lines) and NR patients (blue lines) PT curves are shown for set threshold ranges ($\tau \in [0.1, 0.18]$ for dimension 0 and $\tau \in [0.04, 0.05]$), for the discovery, validation, and both sets combines. P-values and value of the estimators after a t-test comparison are included in Figure S14.

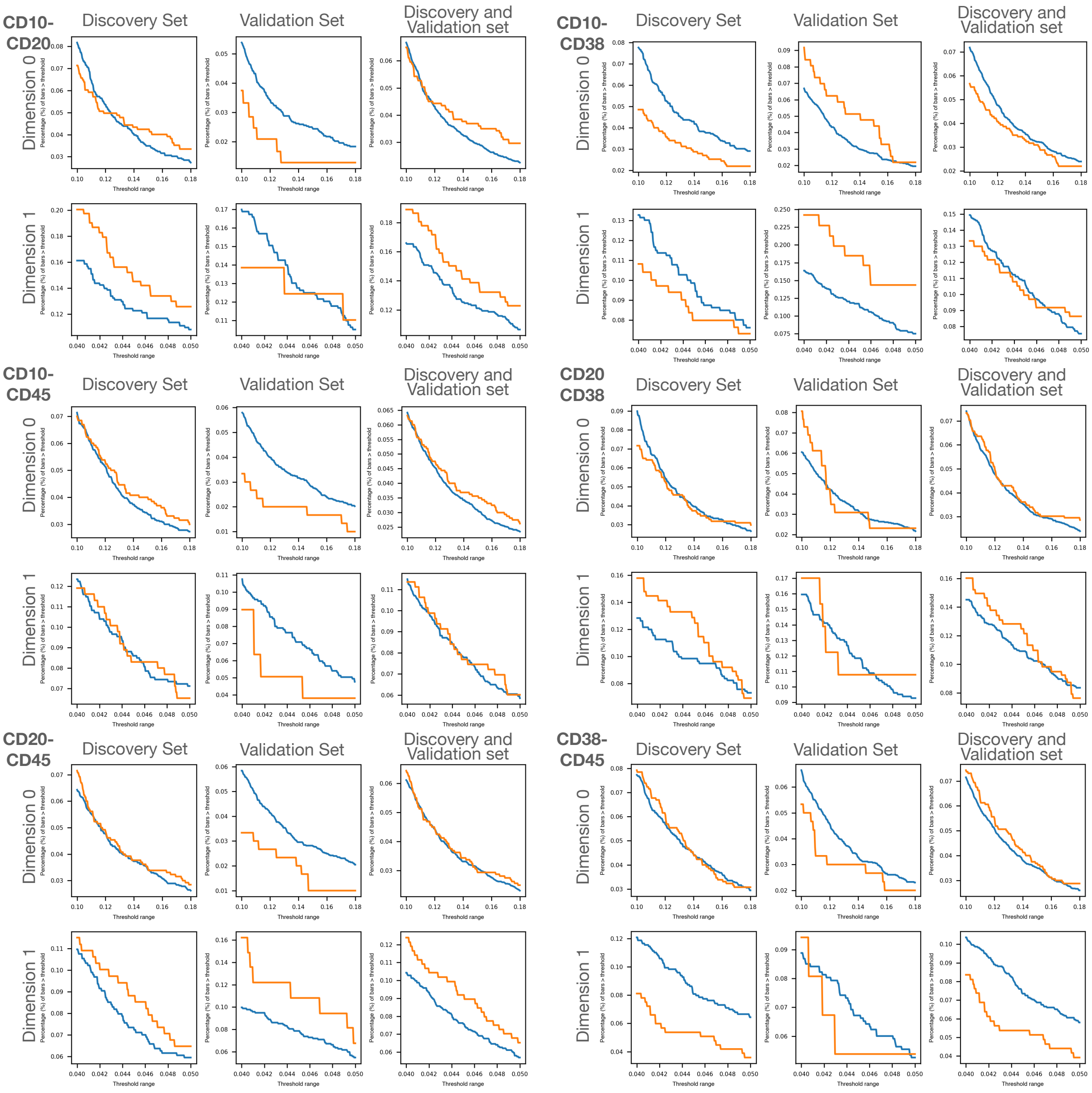

Figure S13: **Difference in the percentage (%) of bars with long persistence in dimension 0 and 1 for the pairwise combination of markers CD10, CD20, CD38 and CD45.** R (orange lines) and NR patients (blue lines) PT curves are shown for set threshold ranges ($\tau \in [0.1, 0.18]$ for dimension 0 and $\tau \in [0.04, 0.05]$), for the discovery, validation, and both sets combines. P-values and value of the estimators after a t-test comparison are included in Figure S15.

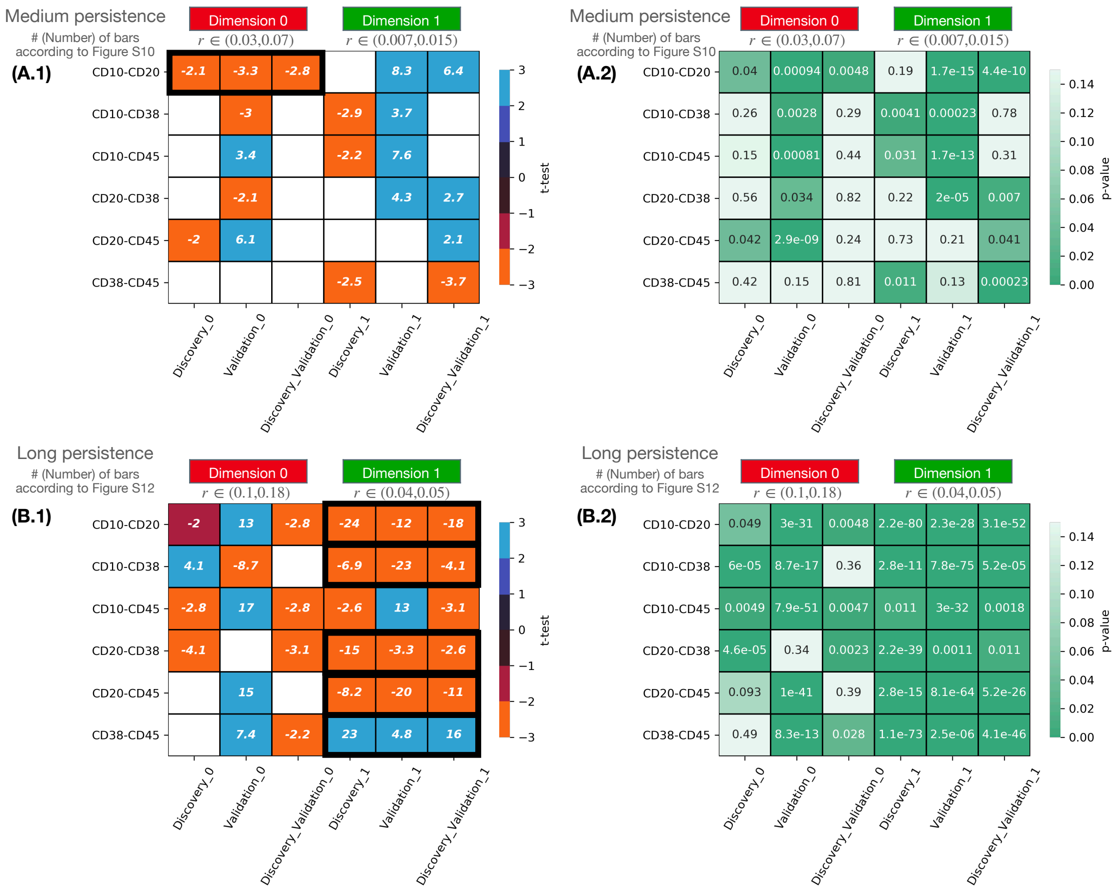

Figure S14: **Difference between NR and R group on number (#) of bars in each patient's PH barcode via persistence threshold curves (PT curves).** This study depended on the dimension (0 and 1) and on the dataset analysed (discovery, validation or both sets). On panel (A.1) and (B.1), we include the value of the estimator after performing a t-test distinguish the PT curve of each cohort of patient, for respectively a number of bars in the range (10,200) (small threshold) and in the range (1,10) (larger threshold). Low values of the estimator (orange) from the t-test correspond to the R group, while high values correspond to the NR group (blue). The corresponding p-values are included in panels (B.1) and (B.2). We highlight in black those difference which are statistically significant and are in agreement between discovery and validation set.

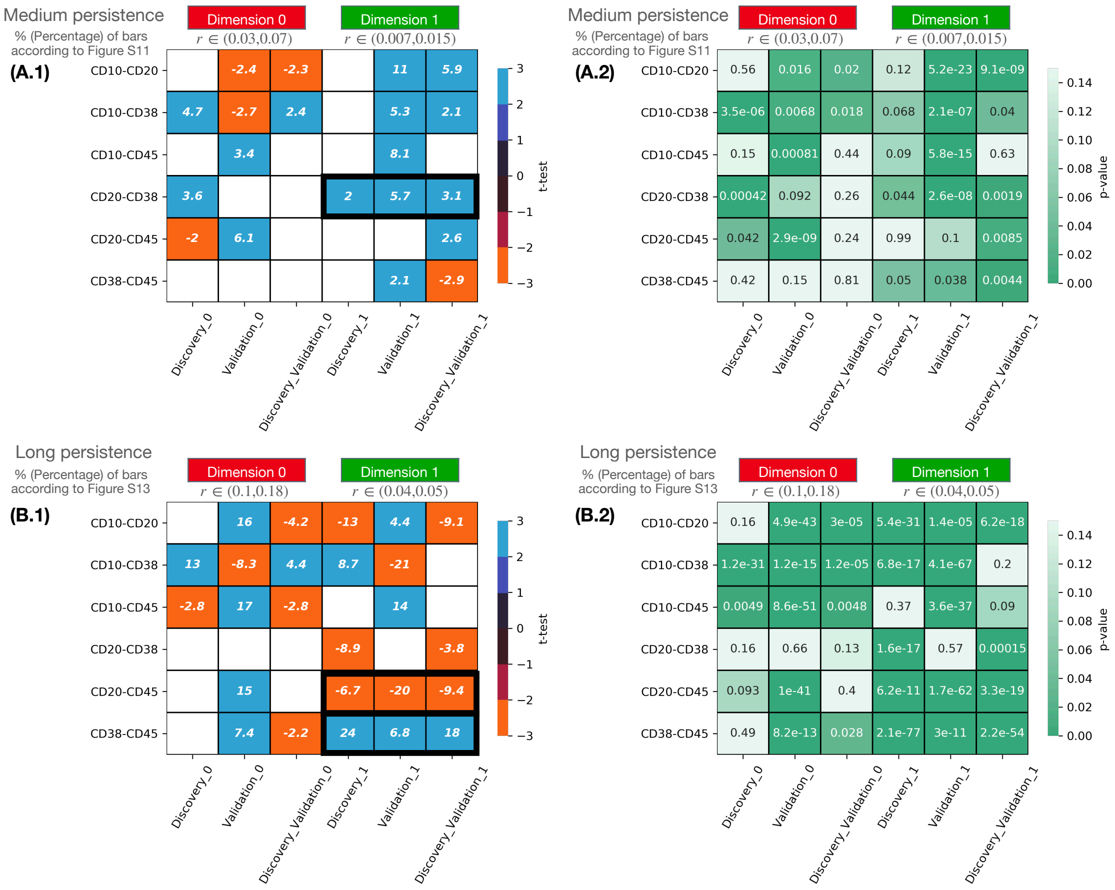

Figure S15: **Difference between NR and R group on percentage (%) of bars over the total number in each patient's PH barcode, via persistence threshold curves (PT curves).** This study depended on the dimension (0 and 1) and on the dataset analysed (discovery, validation or both sets). On panel (A.1) and (B.1), we include the value of the estimator after performing a t-test distinguish the PT curve of each cohort of patient, for respectively a number of bars in the range (10,200) (small threshold) and in the range (1,10) (larger threshold). Low values of the estimator (orange) from the t-test correspond to the R group, while high values correspond to the NR group (blue). The corresponding p-values are included in panels (B.1) and (B.2). We highlight in black those difference which are statistically significant and are in agreement between discovery and validation set.

We compared these results in dimension 0 with a clustering in the four-dimensional space CD10-CD20-CD38-CD45 (see Figures S16, S17 and S18) and in the two-dimensional space of the pair CD10-CD20 (see Figures S19, S20 and S21). This was done by the FlowSom algorithm, obtaining in both analyses fewer connected components in NR patients than in R patients.

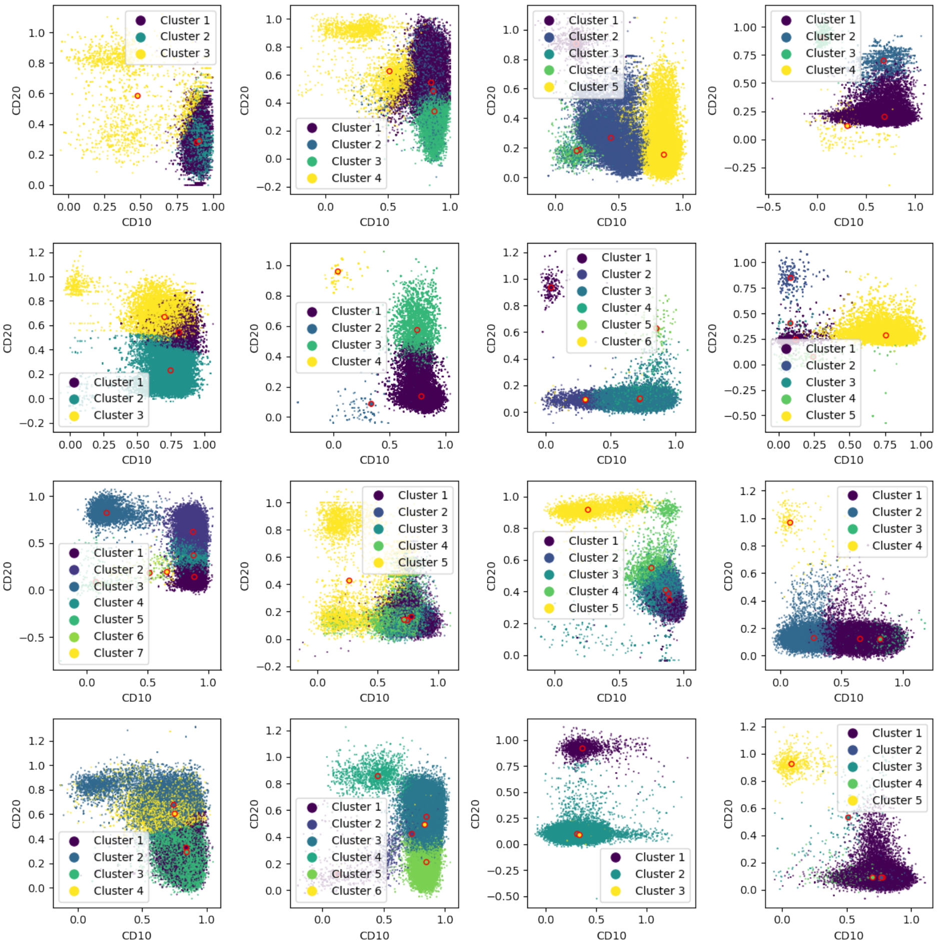

Figure S16: **FlowSom clustering results of CD10-CD20-CD38-CD45 biomarkers, projected in CD10-CD20 for R patients.**

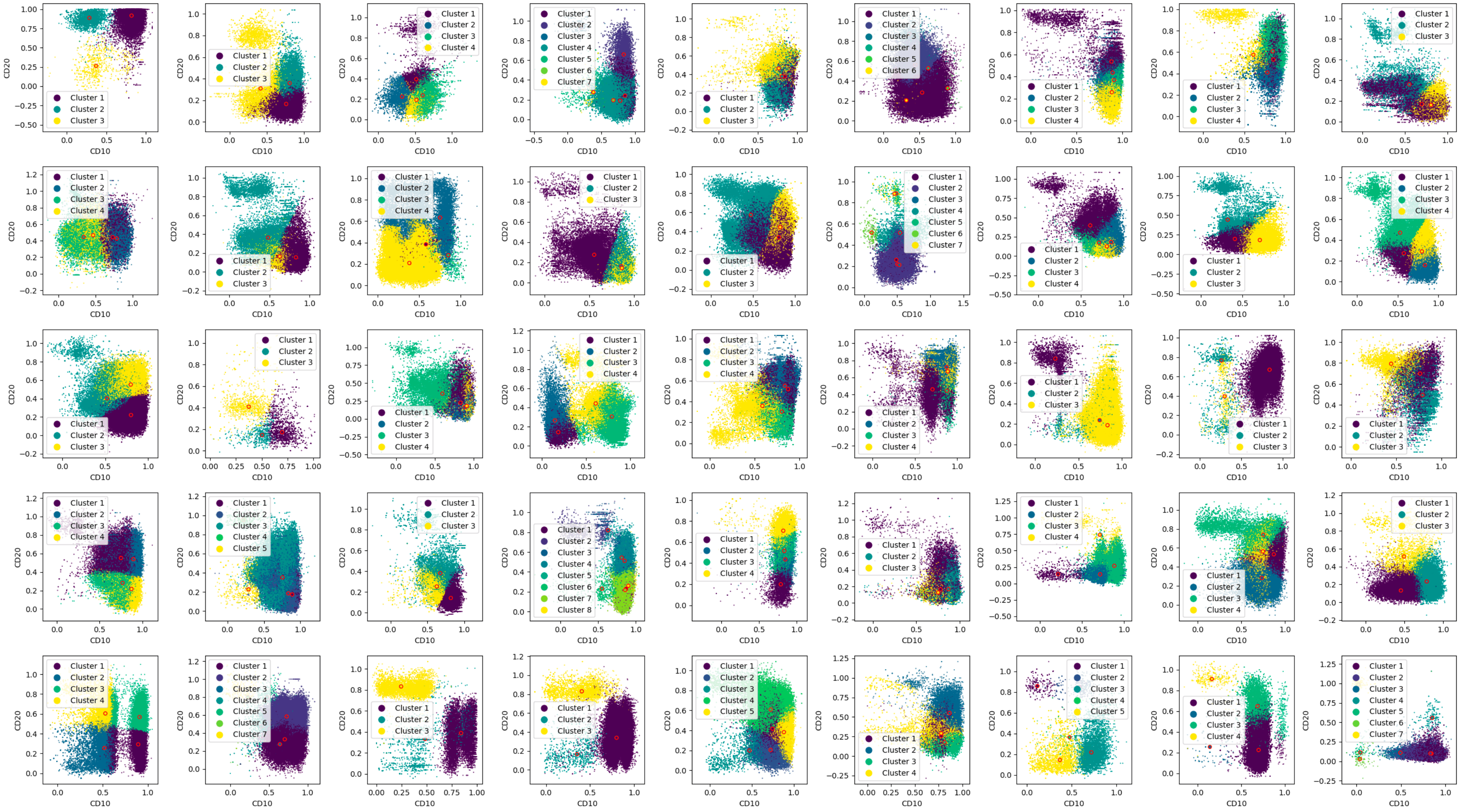

Figure S17: **FlowSom clustering results of CD10-CD20-CD38-CD45 biomarkers, projected in CD10-CD20 for NR patients (I/II)**.

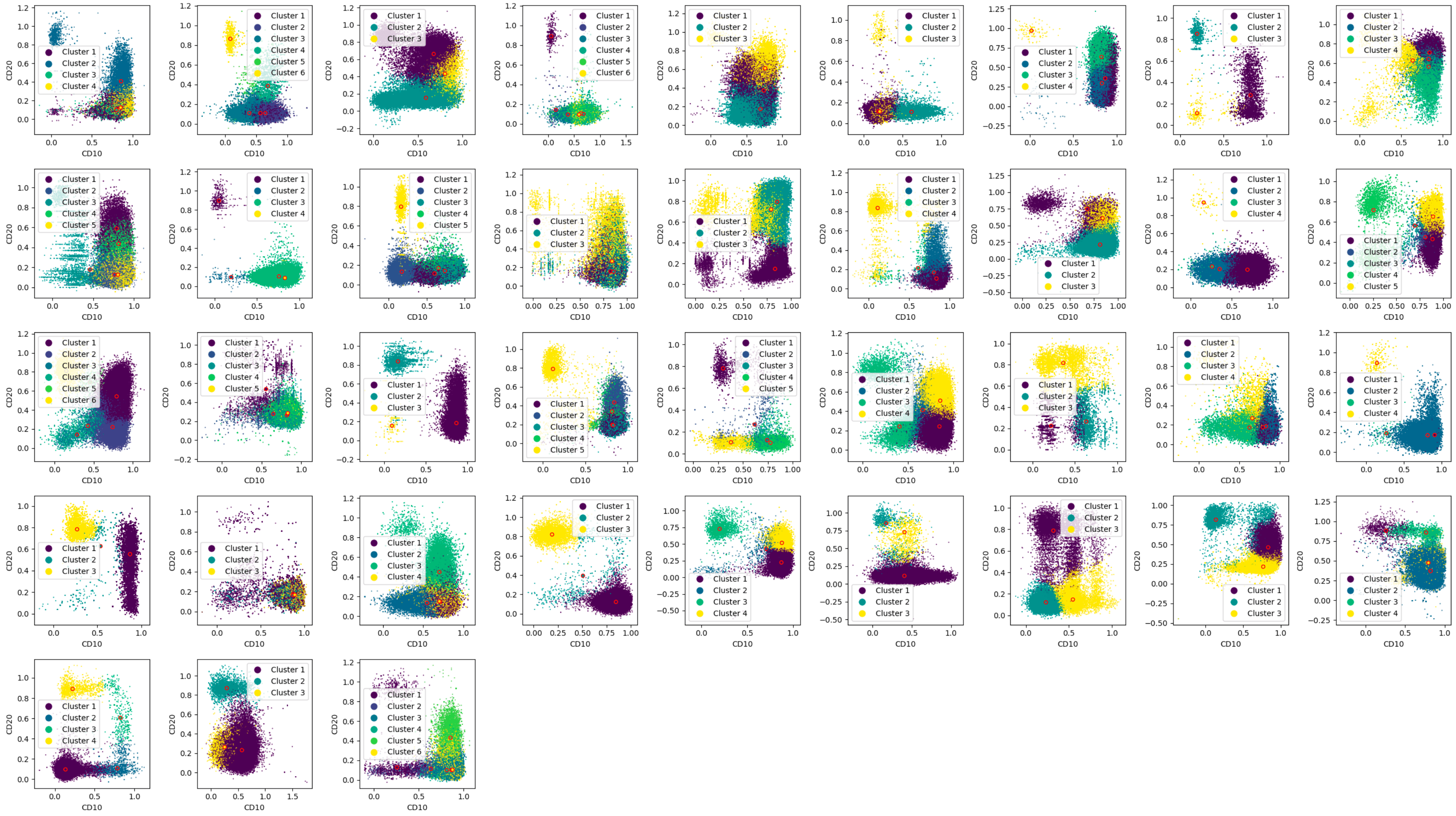

Figure S18: **FlowSom clustering results of CD10-CD20-CD38-CD45 biomarkers, projected in CD10-CD20 for NR patients (II/II).**

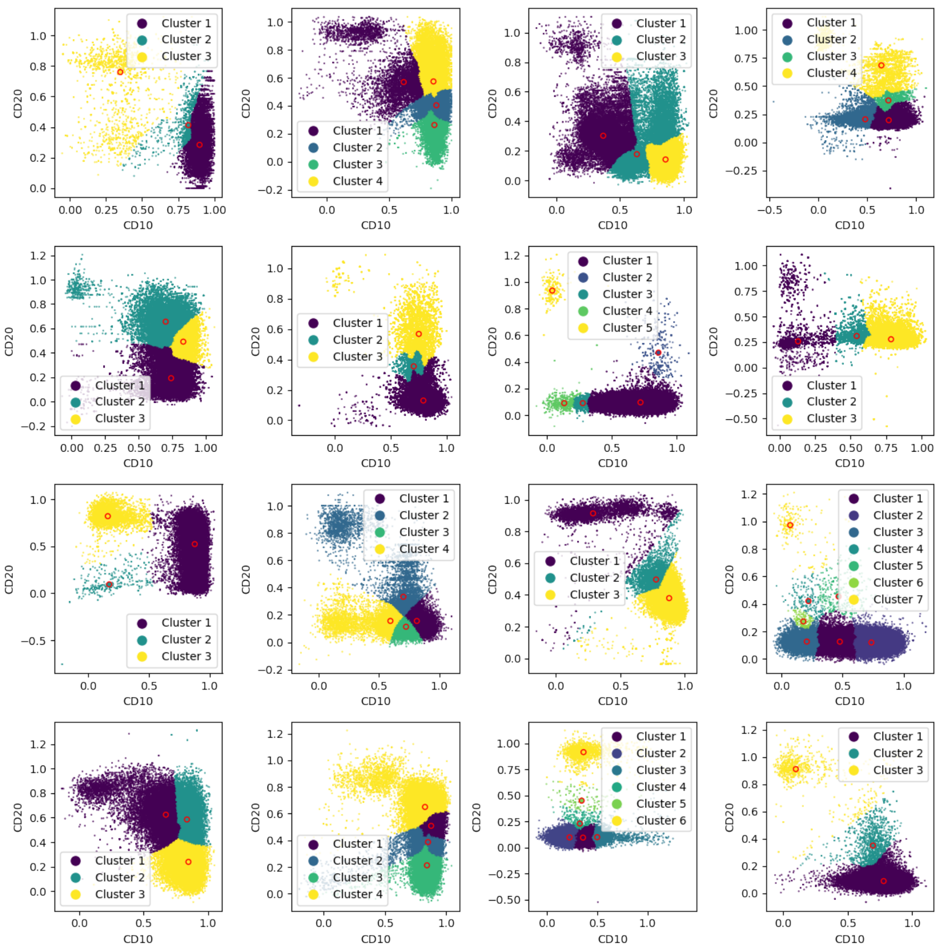

Figure S19: **FlowSom clustering results of CD10-CD20 projection in R patients.**

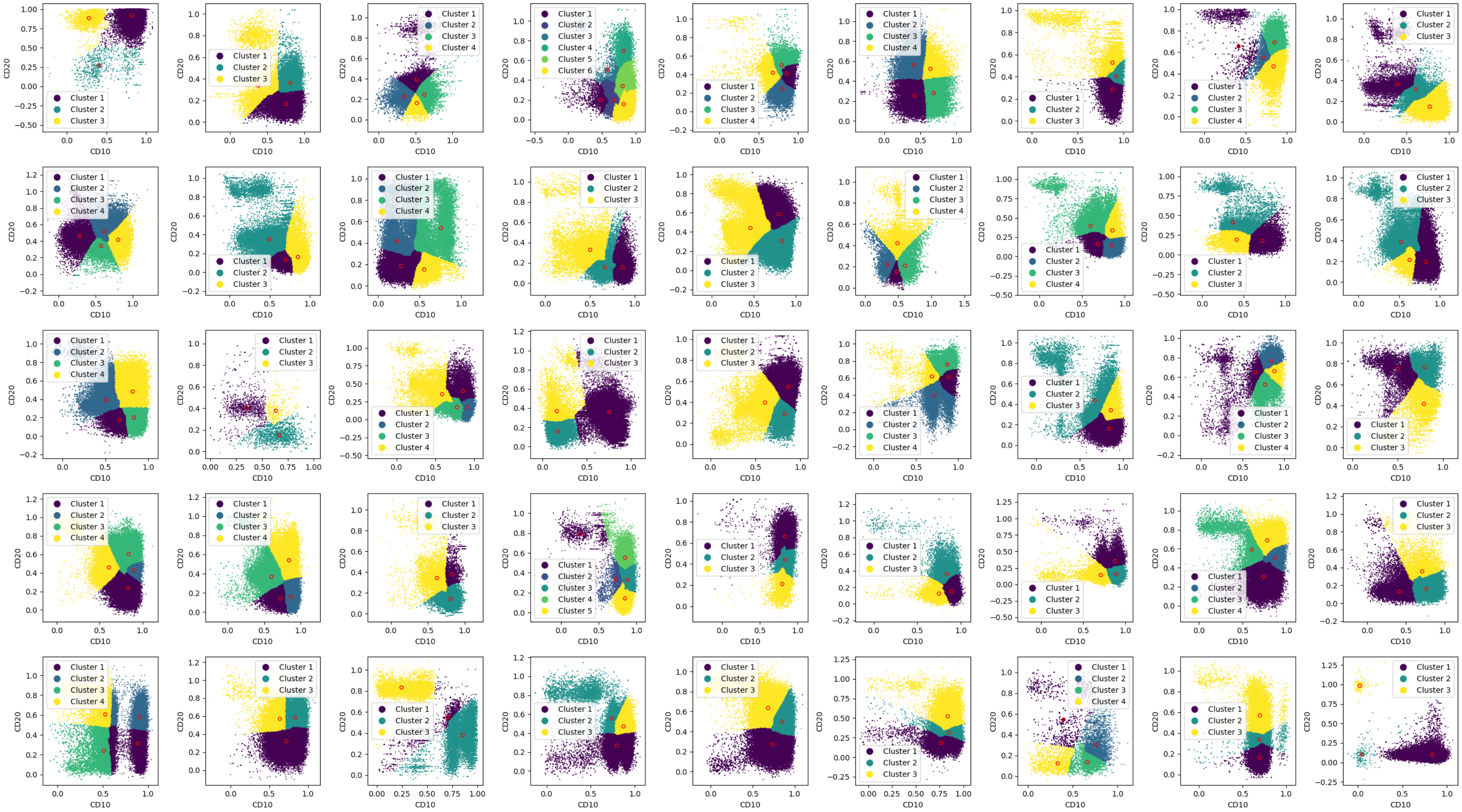

Figure S20: **FlowSom clustering results of CD10-CD20 projection in NR patients (I/II).**.

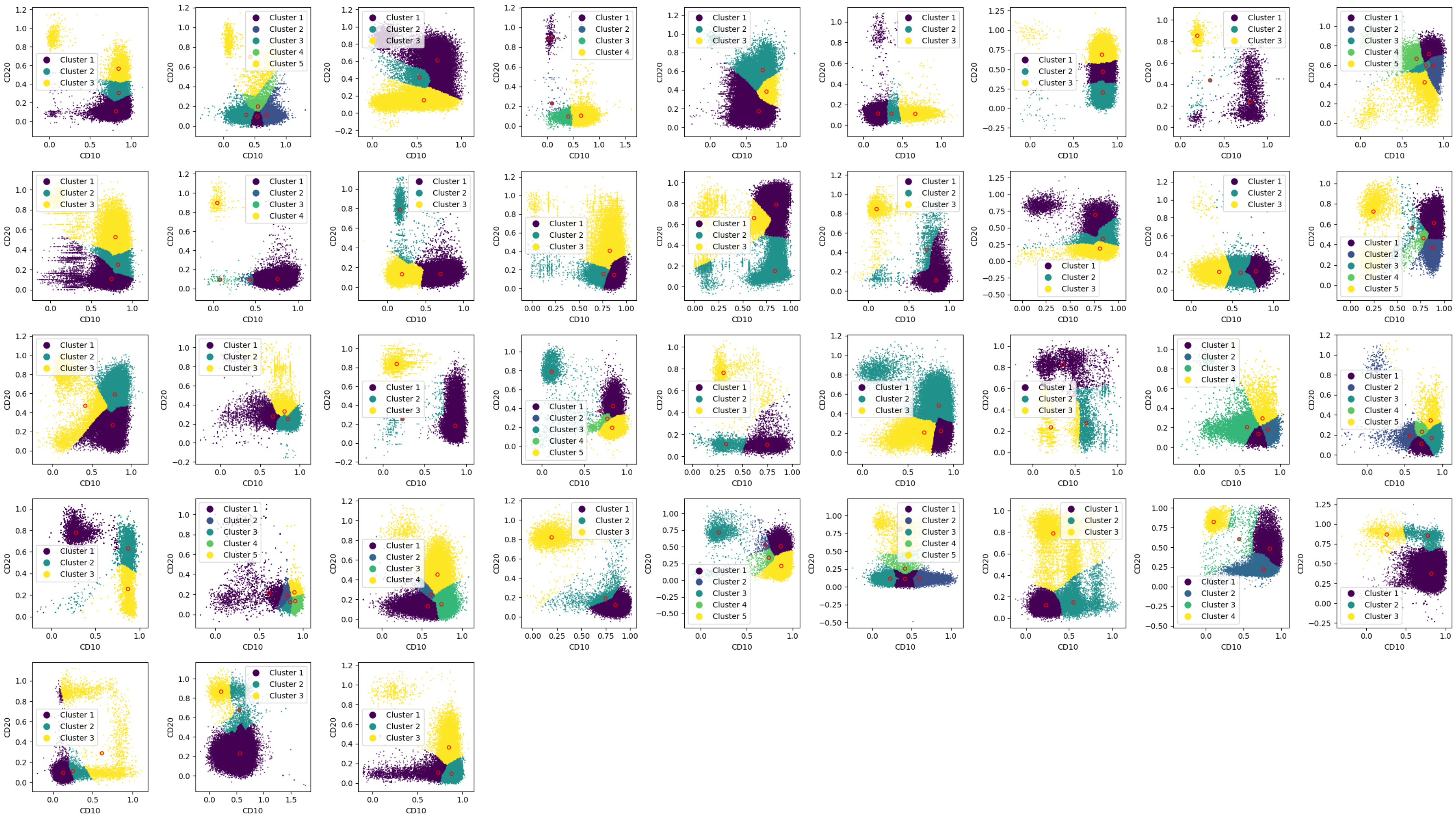

Figure S21: **FlowSom clustering results of CD10-CD20 projection in NR patients (II/II)..**

##### 2.3.c Analysis using persistence images

To classify between R and NR patients, we computed persistence images by transforming the persistence barcodes previously obtained in the four-dimensional space of CD10-CD20-CD38-CD45. Mean persistence images for each patient dataset and results for the most representative and discriminatory areas between both cohorts are included in Figure S22. The complete results depending on the dataset (discovery, validation or both), dimension analysed (0,1 or 2), persistence image grid (50 $\times$50 or 100 $\times$100) and spread of the Gaussian 2D distributions (0.01 or 0.05) for its generation can be found in Figure S23.

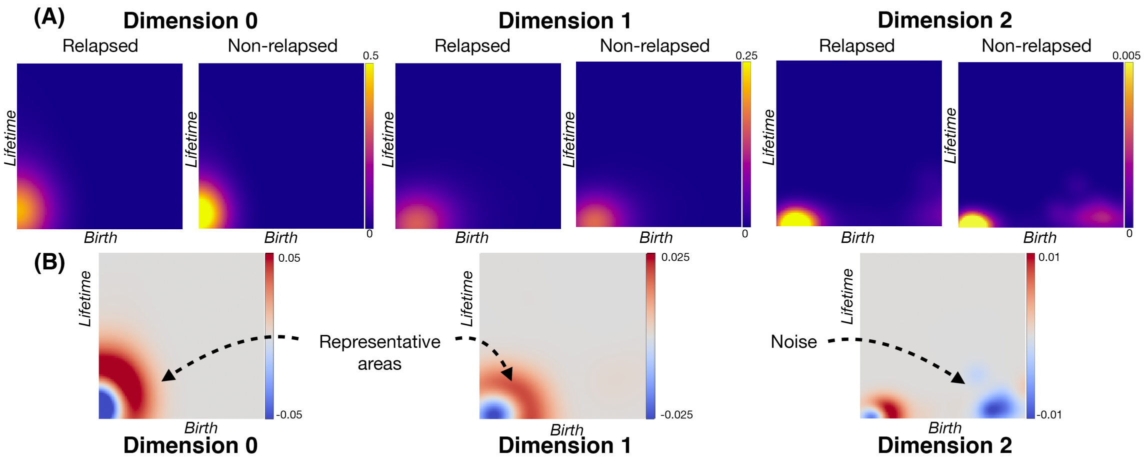

Figure S22: **Persistence images for relapsed and non-relapsed patients.** **(A)** Mean persistence images for each patient set, depending on dimension analysed, with a spread of 0.05 in the 2D Gaussian distribution on a $100 \times 100$ grid. The persistence images obtained are centred around (0,0). The spread influences the width of the regions of high intensity levels. For dimension 0 and 1, non-relapsed patients had more intense profiles than relapsed patients. For dimension 1, relapsed patients had also more spread profiles. Dimension 2 persistence images were also computed, and also presented a data cloud centred around (0,0). However, other data clouds were also produced, indicating either differences between both cohort of patients or noise. **(B)** Difference of representative discriminatory areas after classification between R and NR patients. While dimension 0 focuses on the band around the centre and dimension 1 on the centre itself, both are consistent in the representative areas shown. For dimension 2, other areas are highlighted, and are interpreted as noise, given the classification results shown in Tables S3 and S4.

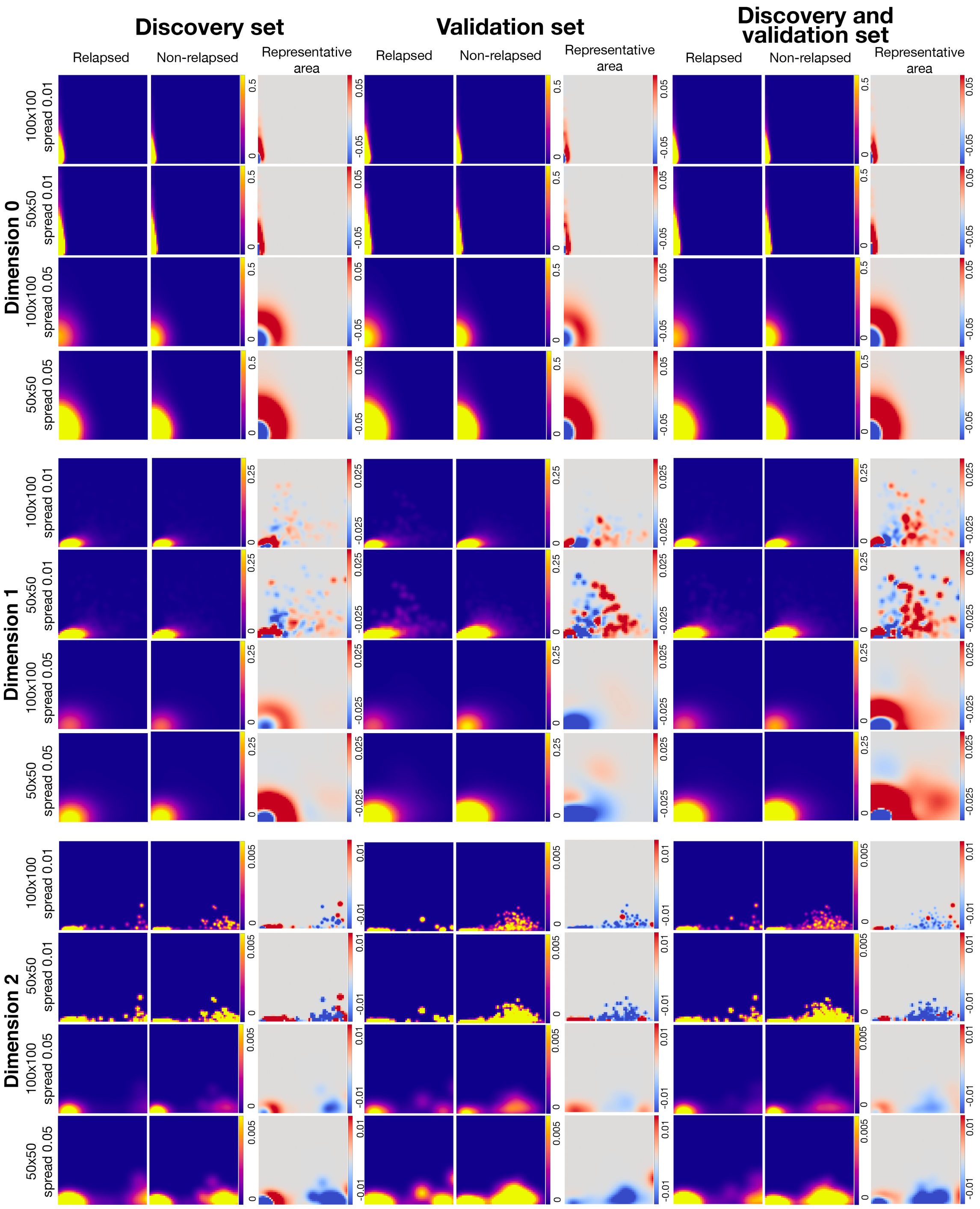

Figure S23: **Mean persistence images for markers CD10, CD20, CD38, CD45.** The results are shown considering the dataset (discovery, validation or both) and dimension analysed (0,1 or 2), which also depends on the choose of persistence image grid (50 ×50 or 100 ×100) and spread of the Gaussian 2D distributions (0.01 or 0.05). The representative area between both cohorts are shown for each dataset. In all images, the x-axis represents the birth of the topological features, while the y-axis is its lifetime or persistence.

##### 2.3.d Classification results

Having obtained the persistence images for R and NR patients, we considered them as input in the form of data matrices for Logistic Regression (LR) and Support Vector Machine (SVM) analyses. We considered multiple analyses: depending on the datasets studied (balanced or not by means of uppersampling), dimensions of the topological features analysed (0, 1 or 2) and markers included (CD10, CD20, CD38, CD45). These results also depend on the dimension, spread and grid used to generate each persistence image, as well as the parameters $C$ and $\gamma$ for the SVM method. The classification results are presented in Tables S3 and S4. We further performed classification by considering all PI images for each patient in each dimension together as input for the SVM method. These results are presented in Table S5.

| **Spread** | **Grid** | **Method** | **Dim. 0** | | | **Dim. 1** | | | **Dim. 2** | | |
|---|---|---|---|---|---|---|---|---|---|---|---|
| | | | **C** | $\gamma$ | **Acc.** | **C** | $\gamma$ | **Acc.** | **C** | $\gamma$ | **Acc.** |
| **0.05** | **100x100** | LR | | | 0.93 | | | 0.93 | | | 0.93 |
| | | SVM | 10 | $10^{-3}$ | 1 | $10^{3}$ | $10^{-4}$ | 1 | $10^{3}$ | 10 | 0.91 |
| | **50x50** | LR | | | 0.97 | | | 0.93 | | | 0.93 |
| | | SVM | 10 | $10^{-3}$ | 1 | $10^{2}$ | $10^{-2}$ | 1 | $10^{4}$ | 1 | 0.89 |
| | **25x25** | LR | | | 1 | | | 0.93 | | | 0.93 |
| | | SVM | 10 | $10^{-4}$ | 1 | $10^{3}$ | $10^{-3}$ | 1 | $10^{7}$ | $10^{-5}$ | 0.91 |
| | **10x10** | LR | | | 1 | | | 0.95 | | | 0.93 |
| | | SVM | 10 | $10^{-5}$ | 1 | $10^{4}$ | $10^{-3}$ | 1 | $10^{2}$ | $10^{-1}$ | 0.89 |
| | **5x5** | LR | | | 1 | | | 0.95 | | | 0.93 |
| | | SVM | 10 | $10^{-5}$ | 1 | $10^{3}$ | $10^{-2}$ | 1 | $10^{4}$ | $10^{-2}$ | 0.93 |
| **0.01** | **100x100** | LR | | | 0.95 | | | 0.91 | | | 0.93 |
| | | SVM | 10 | $10^{-3}$ | 0.95 | $10^{8}$ | $10^{-9}$ | 0.89 | $10^{2}$ | 10 | 0.91 |
| | **50x50** | LR | | | 0.95 | | | 0.89 | | | 0.93 |
| | | SVM | $10^{2}$ | $10^{-4}$ | 0.95 | $10^{9}$ | $10^{-8}$ | 0.78 | 1 | 10 | 0.93 |
| | **25x25** | LR | | | 0.95 | | | 0.87 | | | 0.93 |
| | | SVM | $10^{4}$ | $10^{-6}$ | 0.95 | $10^{4}$ | $10^{-5}$ | 0.89 | $10^{2}$ | $10^{-2}$ | 0.84 |
| | **10x10** | LR | | | 0.91 | | | 0.89 | | | 0.93 |
| | | SVM | $10^{3}$ | $10^{-5}$ | 0.87 | $10^{6}$ | $10^{-6}$ | 0.78 | $10^{2}$ | $10^{-1}$ | 0.93 |
| | **5x5** | LR | | | 0.91 | | | 0.87 | | | 0.91 |
| | | SVM | $10^{4}$ | $10^{-6}$ | 0.89 | $10^{7}$ | $10^{-6}$ | 0.89 | $10^{5}$ | $10^{-2}$ | 0.89 |

Table S3: **Comparison of classification scores of persistence images using the training set and the discovery dataset with testing on the validation dataset.**. Logistic Regression (LR) and Support Vector Machine (SVM) was performed training on the discovery set and testing on the validation set, using biomarkers CD10, CD20, CD38, and CD45. These results depend on the dimension, spread and grid used to generate the Persistence Image, as well as the parameters $C$ and $\gamma$ for the SVM method.

| Spread | Grid | Method | Dim. 0 | | | Dim. 1 | | | Dim. 2 | | |
|---|---|---|---|---|---|---|---|---|---|---|---|
| | | | C | $\gamma$ | Acc. | C | $\gamma$ | Acc. | C | $\gamma$ | Acc. |
| **0.05** | **100x100** | SVM | $10^3$ | $10^{-3}$ | 1 | $10^3$ | $10^{-3}$ | 1 | $10^2$ | $10^2$ | 0.84 |
| | **50x50** | SVM | $10^2$ | $10^{-3}$ | 1 | $10^3$ | $10^{-2}$ | 1 | 10 | $10^2$ | 0.79 |
| | **25x25** | SVM | $10^2$ | $10^{-4}$ | 1 | $10^2$ | $10^{-3}$ | 1 | 10 | 10 | 0.82 |
| | **10x10** | SVM | $10^3$ | $10^{-5}$ | 1 | $10^3$ | $10^{-3}$ | 1 | 10 | 1 | 0.83 |
| | **5x5** | SVM | $10^2$ | $10^{-5}$ | 1 | $10^4$ | $10^{-5}$ | 1 | 10 | 1 | 0.83 |
| **0.01** | **100x100** | SVM | $10^3$ | $10^{-4}$ | 1 | $10^5$ | $10^{-5}$ | 0.97 | $10^2$ | 10 | 0.84 |
| | **50x50** | SVM | $10^3$ | $10^{-5}$ | 1 | $10^5$ | $10^{-5}$ | 0.95 | 10 | 10 | 0.85 |
| | **25x25** | SVM | $10^4$ | $10^{-6}$ | 1 | $10^5$ | $10^{-6}$ | 0.98 | 10 | 10 | 0.84 |
| | **10x10** | SVM | $10^7$ | $10^{-7}$ | 0.81 | 1 | 0.1 | 0.8 | 1 | 10 | 0.81 |
| | **5x5** | SVM | $10^5$ | $10^{-6}$ | 0.8 | 1 | 1 | 0.61 | 10 | 1 | 0.79 |

Table S4: **Comparison of classification scores of persistence images with upper sampling**. Support Vector Machine (SVM) was performed training on the discovery set and testing on the validation set, using biomarkers CD10, CD20, CD38, and CD45. These results depend on the dimension, spread and grid used in the obtention of the Persistence Image, as well as the parameters $C$ and $\gamma$ for the SVM method.

| Spread | Grid | Method | Without Upper sampling | | | With Upper sampling | | |
|---|---|---|---|---|---|---|---|---|
| | | | **C** | $\gamma$ | **Acc.** | **C** | $\gamma$ | **Acc.** |
| 0.05 | **100x100** | SVM | $10^1$ | $10^{-2}$ | 1 | $10^3$ | $10^{-3}$ | 1 |
| | **50x50** | SVM | $10^1$ | $10^{-3}$ | 1 | $10^3$ | $10^{-4}$ | 1 |
| | **25x25** | SVM | $10^1$ | $10^{-3}$ | 1 | $10^3$ | $10^{-5}$ | 1 |
| | **10x10** | SVM | $10^1$ | $10^{-4}$ | 1 | $10^3$ | $10^{-5}$ | 1 |
| | **5x5** | SVM | $10^1$ | $10^{-4}$ | 1 | $10^7$ | $10^{-9}$ | 1 |
| 0.01 | **100x100** | SVM | $10^1$ | $10^{-3}$ | 0.97 | $10^2$ | $10^{-3}$ | 1 |
| | **50x50** | SVM | $10^1$ | $10^{-3}$ | 0.97 | $10^2$ | $10^{-4}$ | 1 |
| | **25x25** | SVM | $10^7$ | $10^{-9}$ | 0.97 | $10^2$ | $10^{-4}$ | 1 |
| | **10x10** | SVM | $10^3$ | $10^{-5}$ | 0.92 | $10^3$ | $10^{-5}$ | 0.93 |
| | **5x5** | SVM | $10^8$ | $10^{-9}$ | 0.9 | $10^4$ | $10^{-7}$ | 0.93 |

Table S5: **Comparison of classification scores of persistence images of dimensions 0, 1 and 2 together as input for the cross-validation**. A classifier based on Support Vector Machine (SVM) was constructed performing cross-validation on the discovery and validation datasets, using the three PIs together obtained in dimension 0, 1 and 2 from PH analysis of biomarkers CD10, CD20, CD38, and CD45. These results depend on the dimension, spread and grid used to generate the Persistence Image, as well as the parameters $C$ and $\gamma$ for the SVM method. We show the results with and without performing uppersampling.

**Data S1. (separate file)**

Random forest analysis of persistence data of pairwise combinations of markers in the whole dataset with oversampling using stratified k-fold.

**Data S2. (separate file)**

Random forest analysis of persistence data of pairwise combinations of markers from the discovery set.

**Data S3. (separate file)**

Random forest analysis of persistence data of pairwise combinations of markers in discovery and validation set.

**Data S4. (separate file)**

Clinical variables of the patients from the discovery and validation set.

| **Dimension** | **Grid** | **Spread** | AUC | Accuracy | Std | Conf. matrix | Recall | F1 | Precision |
|---|---|---|---|---|---|---|---|---|---|
| 0 | 5x5 | 0.01 | 0.70 | 0.78 | 0.05 | [[60 6] [11 0]] | 0.00 | 0.00 | 0.00 |
| | | 0.05 | 1.00 | 1.00 | 0.00 | [[66 0] [ 0 11]] | 1.00 | 1.00 | 1.00 |
| | 10x10 | 0.01 | 0.74 | 0.86 | 0.07 | [[60 6] [ 5 6]] | 0.50 | 0.42 | 0.36 |
| | | 0.05 | 1.00 | 1.00 | 0.00 | [[66 0] [ 0 11]] | 1.00 | 1.00 | 1.00 |
| | 25x25 | 0.01 | 1.00 | 1.00 | 0.00 | [[66 0] [ 0 11]] | 1.00 | 1.00 | 1.00 |
| | | 0.05 | 1.00 | 1.00 | 0.00 | [[66 0] [ 0 11]] | 1.00 | 1.00 | 1.00 |
| | 50x50 | 0.01 | 1.00 | 1.00 | 0.00 | [[66 0] [ 0 11]] | 1.00 | 1.00 | 1.00 |
| | | 0.05 | 1.00 | 1.00 | 0.00 | [[66 0] [ 0 11]] | 1.00 | 1.00 | 1.00 |
| | 100x100 | 0.01 | 1.00 | 1.00 | 0.00 | [[66 0] [ 0 11]] | 1.00 | 1.00 | 1.00 |
| | | 0.05 | 1.00 | 1.00 | 0.00 | [[66 0] [ 0 11]] | 1.00 | 1.00 | 1.00 |
| 1 | 5x5 | 0.01 | 0.62 | 0.84 | 0.00 | [[64 2] [10 1]] | 0.08 | 0.08 | 0.08 |
| | | 0.05 | 1.00 | 1.00 | 0.00 | [[66 0] [ 0 11]] | 1.00 | 1.00 | 1.00 |
| | 10x10 | 0.01 | 0.57 | 0.79 | 0.06 | [[59 7] [ 9 2]] | 0.17 | 0.13 | 0.11 |
| | | 0.05 | 1.00 | 1.00 | 0.00 | [[66 0] [ 0 11]] | 1.00 | 1.00 | 1.00 |
| | 25x25 | 0.01 | 1.00 | 0.97 | 0.04 | [[66 0] [ 2 9]] | 0.75 | 0.78 | 0.83 |
| | | 0.05 | 1.00 | 1.00 | 0.00 | [[66 0] [ 0 11]] | 1.00 | 1.00 | 1.00 |
| | 50x50 | 0.01 | 1.00 | 0.97 | 0.04 | [[66 0] [ 2 9]] | 0.75 | 0.78 | 0.83 |
| | | 0.05 | 1.00 | 1.00 | 0.00 | [[66 0] [ 0 11]] | 1.00 | 1.00 | 1.00 |
| | 100x100 | 0.01 | 1.00 | 0.97 | 0.04 | [[66 0] [ 2 9]] | 0.75 | 0.78 | 0.83 |
| | | 0.05 | 1.00 | 0.97 | 0.06 | [[66 0] [ 2 9]] | 0.83 | 0.83 | 0.83 |
| 2 | 5x5 | 0.01 | 0.78 | 0.78 | 0.10 | [[59 7] [10 1]] | 0.08 | 0.05 | 0.03 |
| | | 0.05 | 0.80 | 0.79 | 0.08 | [[59 7] [ 9 2]] | 0.25 | 0.11 | 75.00 |
| | 10x10 | 0.01 | 0.93 | 0.91 | 0.10 | [[62 4] [ 3 8]] | 0.75 | 0.72 | 0.79 |
| | | 0.05 | 0.93 | 0.93 | 0.08 | [[63 3] [ 2 9]] | 0.83 | 0.79 | 0.81 |
| | 25x25 | 0.01 | 0.88 | 0.88 | 0.10 | [[59 7] [ 2 9]] | 0.83 | 0.69 | 0.65 |
| | | 0.05 | 0.94 | 0.90 | 0.04 | [[66 0] [ 8 3]] | 0.25 | 0.33 | 0.50 |
| | 50x50 | 0.01 | 0.85 | 0.88 | 0.09 | [[60 6] [ 3 8]] | 0.75 | 0.68 | 0.64 |
| | | 0.05 | 0.94 | 0.86 | 0.03 | [[66 0] [11 0]] | 0.00 | 0.00 | 0.00 |
| | 100x100 | 0.01 | 0.86 | 0.83 | 0.08 | [[63 3] [10 1]] | 0.08 | 0.11 | 0.17 |
| | | 0.05 | 0.93 | 0.86 | 0.03 | [[66 0] [11 0]] | 0.00 | 0.00 | 0.00 |

Table S6: **Comparison of full classification scores of persistence images using the whole dataset, without oversampling and using stratified 6-fold validation.**. Support Vector Machine (SVM) was performed on the whole dataset with stratified 6-fold validation, using biomarkers CD10, CD20, CD38, and CD45. These results depend on the dimension, spread and grid used to generate the Persistence Image. The hyperparameters of the SVM classifier were fixed as $C = 10$ and $\gamma = 10^{-3}$.

| Dimension | Grid | Spread | AUC | Accuracy | Std | Conf. matrix | Recall | F1 | Precision |
|---|---|---|---|---|---|---|---|---|---|
| 0 | 5x5 | 0.01 | 0.86 | 0.8 | 0.07 | [[49 17] [ 9 53]] | 0.85 | 0.8 | 0.76 |
| | | 0.05 | 1.0 | 1 | 0.0 | [[66 0] [ 0 62]] | 1.0 | 1.0 | 1.0 |
| | 10x10 | 0.01 | 0.94 | 0.91 | 0.07 | [[58 8] [ 3 59]] | 0.95 | 0.92 | 0.89 |
| | | 0.05 | 1.0 | 1.0 | 0.0 | [[66 0] [ 0 62]] | 1.0 | 1.0 | 1.0 |
| | 25x25 | 0.01 | 1.0 | 1.0 | 0.0 | [[66 0] [ 0 62]] | 1.0 | 1.0 | 1.0 |
| | | 0.05 | 1.0 | 1.0 | 0.0 | [[66 0] [ 0 62]] | 1.0 | 1.0 | 1.0 |
| | 50x50 | 0.01 | 1.0 | 1.0 | 0.0 | [[66 0] [ 0 62]] | 1.0 | 1.0 | 1.0 |
| | | 0.05 | 1.0 | 1.0 | 0.0 | [[66 0] [ 0 62]] | 1.0 | 1.0 | 1.0 |
| | 100x100 | 0.01 | 1.0 | 1.0 | 0.0 | [[66 0] [ 0 62]] | 1.0 | 1.0 | 1.0 |
| | | 0.05 | 1.0 | 1.0 | 0.0 | [[66 0] [ 0 62]] | 1.0 | 1.0 | 1.0 |
| 1 | 5x5 | 0.01 | 0.67 | 0.69 | 0.06 | [[40 26] [14 48]] | 0.77 | 0.7 | 0.67 |
| | | 0.05 | 1.0 | 1.0 | 0.0 | [[66 0] [ 0 62]] | 1.0 | 1.0 | 1.0 |
| | 10x10 | 0.01 | 0.93 | 0.94 | 0.06 | [[58 8] [ 0 62]] | 1.0 | 0.94 | 0.9 |
| | | 0.05 | 1.0 | 1.0 | 0.0 | [[66 0] [ 0 62]] | 1.0 | 1.0 | 1.0 |
| | 25x25 | 0.01 | 1.0 | 1.0 | 0.0 | [[66 0] [ 0 62]] | 1.0 | 1.0 | 1.0 |
| | | 0.05 | 1.0 | 1.0 | 0.0 | [[66 0] [ 0 62]] | 1.0 | 1.0 | 1.0 |
| | 50x50 | 0.01 | 1.0 | 1.0 | 0.0 | [[66 0] [ 0 62]] | 1.0 | 1.0 | 1.0 |
| | | 0.05 | 1.0 | 1.0 | 0.0 | [[66 0] [ 0 62]] | 1.0 | 1.0 | 1.0 |
| | 100x100 | 0.01 | 1.0 | 1.0 | 0.0 | [[66 0] [ 0 62]] | 1.0 | 1.0 | 1.0 |
| | | 0.05 | 1.0 | 1.0 | 0.0 | [[66 0] [ 0 62]] | 1.0 | 1.0 | 1.0 |
| 2 | 5x5 | 0.01 | 0.88 | 0.84 | 0.07 | [[55 11] [ 9 53]] | 0.86 | 0.84 | 0.83 |
| | | 0.05 | 0.92 | 0.87 | 0.06 | [[55 11] [ 6 56]] | 0.90 | 0.86 | 0.84 |
| | 10x10 | 0.01 | 0.94 | 0.85 | 0.05 | [[53 13] [ 6 56]] | 0.90 | 0.86 | 0.82 |
| | | 0.05 | 0.96 | 0.92 | 0.02 | [[60 6] [ 4 58]] | 0.93 | 0.92 | 0.91 |
| | 25x25 | 0.01 | 0.9 | 0.89 | 0.07 | [[55 11] [ 3 59]] | 0.95 | 0.89 | 0.85 |
| | | 0.05 | 0.97 | 0.93 | 0.03 | [[61 5] [ 4 58]] | 0.93 | 0.93 | 0.93 |
| | 50x50 | 0.01 | 0.96 | 0.9 | 0.07 | [[56 10] [ 3 59]] | 0.95 | 0.9 | 0.86 |
| | | 0.05 | 0.97 | 0.87 | 0.05 | [[53 13] [ 4 58]] | 0.93 | 0.87 | 0.83 |
| | 100x100 | 0.01 | 0.95 | 0.89 | 0.08 | [[55 11] [ 3 59]] | 0.95 | 0.9 | 0.85 |
| | | 0.05 | 0.95 | 0.8 | 0.11 | [[44 22] [ 4 58]] | 0.93 | 0.82 | 0.75 |

Table S7: **Comparison of full classification scores of persistence images using the whole dataset, with oversampling and stratified 6-fold validation.**. Support Vector Machine (SVM) was performed on the whole dataset with stratified 6-fold validation, using biomarkers CD10, CD20, CD38, and CD45. These results depend on the dimension, spread and grid used to generate the Persistence Image. The hyperparameters of the SVM classifier were fixed as $C = 10$ and $\gamma = 10^{-3}$.

| **Dimension** | **Grid** | **Spread** | AUC | Accuracy | Std | Conf. matrix | Recall | F1 | Precision |
|---|---|---|---|---|---|---|---|---|---|
| 0 | 5x5 | 0.01 | 0.75 | 0.81 | 0.10 | [[54 5] [ 8 1]] | 0.08 | 0.06 | 0.04 |
| | | 0.05 | 1.00 | 1.00 | 0.00 | [[59 0] [ 0 9]] | 1.00 | 1.00 | 1.00 |
| | 10x10 | 0.01 | 0.86 | 0.85 | 0.08 | [[54 5] [ 5 4]] | 0.50 | 0.43 | 0.39 |
| | | 0.05 | 1.00 | 1.00 | 0.00 | [[59 0] [ 0 9]] | 1.00 | 1.00 | 1.00 |
| | 25x25 | 0.01 | 1.00 | 0.99 | 0.03 | [[59 0] [ 1 8]] | 0.92 | 0.94 | 1.00 |
| | | 0.05 | 1.00 | 1.00 | 0.00 | [[59 0] [ 0 9]] | 1.00 | 1.00 | 1.00 |
| | 50x50 | 0.01 | 1.00 | 0.99 | 0.03 | [[59 0] [ 1 8]] | 0.92 | 0.94 | 1.00 |
| | | 0.05 | 1.00 | 1.00 | 0.00 | [[59 0] [ 0 9]] | 1.00 | 1.00 | 1.00 |
| | 100x100 | 0.01 | 1.00 | 0.99 | 0.03 | [[59 0] [ 1 8]] | 0.92 | 0.94 | 1.00 |
| | | 0.05 | 1.00 | 1.00 | 0.00 | [[59 0] [ 0 9]] | 1.00 | 1.00 | 1.00 |
| 1 | 5x5 | 0.01 | 0.59 | 0.80 | 0.07 | [[54 5] [ 9 0]] | 0.00 | 0.00 | 0.00 |
| | | 0.05 | 1.00 | 1.00 | 0.00 | [[59 0] [ 0 9]] | 1.00 | 1.00 | 1.00 |
| | 10x10 | 0.01 | 0.64 | 0.82 | 0.11 | [[52 7] [ 5 4]] | 0.50 | 0.44 | 0.42 |
| | | 0.05 | 1.00 | 1.00 | 0.00 | [[59 0] [ 0 9]] | 1.00 | 1.00 | 1.00 |
| | 25x25 | 0.01 | 1.00 | 1.00 | 0.00 | [[59 0] [ 0 9]] | 1.00 | 1.00 | 1.00 |
| | | 0.05 | 1.00 | 1.00 | 0.00 | [[59 0] [ 0 9]] | 1.00 | 1.00 | 1.00 |
| | 50x50 | 0.01 | 1.00 | 1.00 | 0.00 | [[59 0] [ 0 9]] | 1.00 | 1.00 | 1.00 |
| | | 0.05 | 1.00 | 1.00 | 0.00 | [[59 0] [ 0 9]] | 1.00 | 1.00 | 1.00 |
| | 100x100 | 0.01 | 1.00 | 1.00 | 0.00 | [[59 0] [ 0 9]] | 1.00 | 1.00 | 1.00 |
| | | 0.05 | 1.00 | 0.99 | 0.03 | [[59 0] [ 1 8]] | 0.92 | 0.94 | 1.00 |
| 2 | 5x5 | 0.01 | 0.80 | 0.85 | 0.04 | [[53 6] [ 4 5]] | 0.58 | 0.41 | 0.33 |
| | | 0.05 | 0.92 | 0.82 | 0.05 | [[53 6] [ 6 3]] | 0.33 | 0.28 | 0.25 |
| | 10x10 | 0.01 | 0.81 | 0.87 | 0.10 | [[54 5] [ 4 5]] | 0.58 | 0.57 | 0.64 |
| | | 0.05 | 0.87 | 0.90 | 0.08 | [[55 4] [ 3 6]] | 0.67 | 0.61 | 0.64 |
| | 25x25 | 0.01 | 0.74 | 0.82 | 0.11 | [[51 8] [ 4 5]] | 0.50 | 0.37 | 0.33 |
| | | 0.05 | 0.93 | 0.84 | 0.06 | [[57 2] [ 9 0]] | 0.00 | 0.00 | 0.00 |
| | 50x50 | 0.01 | 0.80 | 0.84 | 0.13 | [[52 7] [ 4 5]] | 0.50 | 0.36 | 0.31 |
| | | 0.05 | 0.93 | 0.87 | 0.04 | [[59 0] [ 9 0]] | 0.00 | 0.00 | 0.00 |
| | 100x100 | 0.01 | 775.00 | 0.84 | 0.06 | [[57 2] [ 9 0]] | 0.00 | 0.00 | 0.00 |
| | | 0.05 | 0.93 | 0.87 | 0.04 | [[59 0] [ 9 0]] | 0.00 | 0.00 | 0.00 |

Table S8: **Comparison of full classification scores of persistence images using the low-intermediate patients dataset, without oversampling and using stratified 6-fold validation.**. Support Vector Machine (SVM) was performed on the whole dataset with stratified 6-fold validation, using biomarkers CD10, CD20, CD38, and CD45. These results depend on the dimension, spread and grid used to generate the Persistence Image. The hyperparameters of the SVM classifier were fixed as $C = 10$ and $\gamma = 10^{-3}$.
